# Center-surround processing in psychosis

**DOI:** 10.1101/2025.08.15.25333221

**Authors:** Samantha A. Montoya, Hannah R. Moser, Rohit S. Kamath, Li Shen Chong, Andrea N. Grant, Małgorzata Marjańska, Scott R. Sponheim, Stephen A. Engel, Cheryl A. Olman, Michael-Paul Schallmo

**Affiliations:** Graduate Program in Neuroscience, University of Minnesota, Minneapolis, MN; Department of Psychiatry and Behavioral Sciences, University of Minnesota, Minneapolis, MN; Center for Magnetic Resonance Research, Department of Radiology, University of Minnesota, Minneapolis, MN; Veterans Affairs Medical Center, Minneapolis, MN; Department of Psychology, University of Minnesota, Minneapolis, MN

## Abstract

**Purpose:** People with psychotic psychopathology (PwPP) often experience subtle variations in visual perception, which can be quantified experimentally. In the contrast surround suppression illusion, a central pattern appears to have lower contrast in the presence of a surrounding pattern. PwPP typically show weaker contrast suppression from the surround than controls, but the mechanisms underlying this difference are still poorly understood.

**Methods:** We assessed perceptual and neural surround suppression in 38 controls, 44 first-degree biological relatives of PwPP, and 64 PwPP as part of the Psychosis Human Connectome Project. To better understand neural mechanisms contributing to diminished surround suppression we quantified contrast discrimination thresholds and examined 7 tesla fMRI responses in the lateral geniculate nucleus (LGN), primary visual cortex (V1), and lateral occipital complex (LOC). Additionally, we measured the concentration of γ-aminobutyric acid (GABA; an inhibitory neurotransmitter) in occipital cortex using 7 T MR spectroscopy.

**Results:** Responses in LOC showed the expected effect of weaker surround suppression in PwPP and relatives versus controls. However, in V1 we found no differences in surround suppression strength between controls, relatives, and PwPP. Additionally, we saw no behavioral evidence for reduced surround suppression in PwPP. Suppression metrics were not significantly correlated with occipital GABA levels or symptom measures. Multi-voxel pattern analysis of V1 fMRI responses revealed a group difference in decoding Surround vs. No Surround, with a trend toward lower accuracy in PwPP vs. controls.

**Conclusion:** Our results suggest subtle differences in visual center-surround processing among people with schizophrenia. Possible explanations for the discrepancy with previous findings include differences in task design and the deployment of spatial attention across groups. Poorer decoding of center vs. surround may suggest neural representations of spatial context in V1 are less reliable in PwPP.

## 1 Introduction

The biological bases of psychotic disorders (e.g., schizophrenia) are poorly understood. One promising avenue for improving our understanding of psychosis is through studying the visual system in people with psychotic psychopathology (PwPP) compared to controls. Firstly, the neural bases of visual perception are relatively well-studied in healthy adults and animal models, providing a good understanding of neurotypical visual systems for comparison. Secondly, visual perception is often altered in people with psychotic disorders. In addition to overt hallucinations, PwPP also experience altered perception of external stimuli. These subtler distortions in visual perception have been observed in a variety of visual tasks (for reviews, see Butler et al., 2008; King et al., 2017; Klein et al., 2020; Notredame et al., 2014; Phillips & Silverstein, 2013; Yoon et al., 2013).

One visual task that has elicited differences between PwPP and controls is contrast surround suppression. Contrast surround suppression is an illusion in which perception of a central target tends to be suppressed by a similar surrounding pattern. Neural responses to central targets are also reduced by the presence of similar patterns presented outside of the classical receptive field (Angelucci & Bressloff, 2006; Cavanaugh et al., 2002a; Nurminen & Angelucci, 2014; Webb et al., 2005). Surround suppression is thought to aid in detecting the boundaries of objects by suppressing responses to contiguous patterns, and is observed in many species including mice, cats, and nonhuman primates (Cavanaugh et al., 2002a; Chisum et al., 2003; Jones et al., 2001, 2002; Levitt & Lund, 2002; Li et al., 2024; Ozeki et al., 2004; Self et al., 2014; Shushruth et al., 2009; Vaiceliunaite et al., 2013; Van den Bergh et al., 2010). Interestingly, this illusion is generally weaker in PwPP, such that PwPP are typically more accurate than controls at judging the contrast of a target stimulus in the presence of a surround (Kéri et al., 2005; Must et al., 2004; Serrano-Pedraza et al., 2014) although there are some conflicting reports (Barch et al., 2012; Chen et al., 2008). While reduced surround suppression has been observed using behavioral tasks in people with schizophrenia and bipolar disorder, relatively little is known about the neural mechanisms underlying these group differences. Elucidating such mechanisms may aid in understanding the pathophysiology of psychotic disorders and how they affect the visual system.

Possible mechanisms include reduced neural suppression in early visual areas among PwPP (e.g., primary visual cortex; V1), as suggested by a couple of studies using functional MRI (fMRI; Anderson et al., 2017; Seymour et al., 2013). At the neurochemical level, studies in animal models have suggested that surround suppression may depend on inhibitory (i.e., γ-aminobutyric acid; GABA) and / or excitatory (i.e., glutamate) mechanisms (Adesnik et al., 2012; Haider et al., 2010; Kirchberger et al., 2023; Liu et al., 2018; Ma et al., 2010; Nienborg et al., 2013; Ozeki et al., 2004, 2009; Sato et al., 2016). In humans, some studies using MR spectroscopy have suggested a relationship between lower GABA levels in visual cortex and weaker surround suppression in both healthy adults (Cook et al., 2016), and people with schizophrenia (Yoon et al., 2010), though others have not observed significant relationships (Schallmo et al., 2018; Schallmo et al., 2020).

It is not clear to what extent genetic factors may contribute to altered surround suppression in PwPP. Genetics play a large role in the development of psychotic disorders (Cardno et al., 1999; Cardno & Owen, 2014), but the specific role of genetics in visual disturbances is poorly understood. One approach for investigating this is to study visual functioning among first-degree biological relatives of PwPP (e.g., parents, siblings, children), who share on average 50% of their genes with PwPP, but do not have a psychotic disorder themselves. Previous studies from our group investigating visual context processing among relatives of PwPP have shown modest (Pokorny et al., 2023), or no differences (Schallmo et al., 2013, 2015) versus healthy controls.

Here, we sought to elucidate the mechanisms of altered surround suppression in PwPP as part of the Psychosis Human Connectome Project—a large, transdiagnostic study with clinical, behavioral, and neuroimaging data (for more information, see Demro et al., 2021; Schallmo et al., 2023). To investigate the perceptual and neural bases of visual dysfunction in PwPP, we measured surround suppression using both behavioral psychophysics and 7 T functional MRI. We also measured neurochemical concentrations (e.g., GABA, glutamate) in the occipital lobe using 7 T MR spectroscopy, and symptoms levels via clinical assessments. We studied three groups of participants: PwPP, first-degree biological relatives, and controls, providing the largest, publicly available, multi-modal dataset of center-surround suppression in PwPP to date. We expected that behavioral and neural (i.e., fMRI responses in V1) surround suppression would be reduced in PwPP compared to controls, and that this might correlate with differences in the levels of occipital neurochemicals, and / or with measures of psychiatric symptoms. Instead, our results suggest a more nuanced view of surround suppression in psychosis.

## 2 Methods

### 2.1 Participants

#### Demographics

As part of the Psychosis Human Connectome Project (HCP), PwPP (n = 64), first-degree biological relatives of PwPP (n = 44), and controls with no family history of psychosis (n = 38) were recruited through the Veterans Affairs Center, Fairview Riverside Hospital in Minneapolis, and Psychiatry Department at the University of Minnesota. A total of 146 participants between the ages of 18 and 60 years participated in the study, though not all participants completed all experiments (see Supplemental Table 1). Data on sex assigned at birth, age, estimated IQ, years of education, and race can be found in Table 1. A subset of participants (10 controls, 39 PwPP) returned for a repeat scanning session at least 1 month after their initial session. For this reason, the number of data sets is larger than the number of unique participants.

**Table 1.** Participant Demographics. Data are presented as mean (standard deviation), unless otherwise noted. Participant diagnoses were made using the Structured Clinical Interview for DSM-IV-TR disorders (SCID; First, 2015). Estimated IQ was determined with the Wechsler Adult Intelligence Scale (WAIS-IV; Petermann & Wechsler, 2012). Racial and ethnic designations (as defined by the National Institute of Health) are abbreviated as follows: A = Asian or Pacific Islander, B = Black (not of Hispanic origin), H = Hispanic, N = Native American or Alaskan Native, W = White (not of Hispanic origin), M = More than 1 race or ethnicity, or other. BACS = Brief Assessment of Cognition in Schizophrenia, Z-score (Keefe et al., 2004), BPRS = Brief Psychiatric Rating Scale Total Score (Ventura et al., 2000), SPQ = Schizotypal Personality Questionnaire (Raine, 1991). Data collected at repeat scans are not included in this table. The test statistics and p values for differences across all 3 groups are shown in the statistics column. Non-parametric statistics (Kruskal-Wallis tests, X^2^ values) were used in place of ANOVAs in cases where data were not normally distributed and / or variance across groups was not homogeneous.

|  | Control (38) | Relative (44) | PwPP (64) | Statistics |
| --- | --- | --- | --- | --- |
| Sex (% female) | 50.0 | 68.2 | 48.4 | $\chi^2_{(2)} = 4.58, p = 0.10$ |
| Age (years) | 39.4 (11.5) | 43.5 (13.0) | 38.1 (11.7) | $F_{(2, 143)} = 2.64, p = 0.075$ |
| Estimated IQ | 107.8 (9.5) | 103.9 (9.8) | 97.5 (10.7) | $F_{(2, 143)} = 13.4, p = 5 \times 10^{-6}$ |
| Education (years) | 16.3 (2.0) | 15.5 (2.1) | 14.2 (2.3) | $F_{(2, 143)} = 11.4, p = 3 \times 10^{-5}$ |
| Visual acuity (decimal fraction) | 0.92 (0.28) | 0.97 (0.31) | 0.95 (0.55) | $F_{(2, 108)} = 0.07, p = 0.9$ |
| Race / ethnicity (%; A / B / H / N / W / M) | 2.6 / 5.3 / 0 / 0 / 89.5 / 2.6 | 2.3 / 4.6 / 0 / 0 / 88.6 / 4.6 | 4.7 / 15.6 / 3.1 / 0 / 73.4 / 3.1 | - |
| BACS | 0.42 (0.53) | 0.01 (0.74) | -0.67 (0.69) | $F_{(2, 142)} = 33.96, p = 9 \times 10^{-13}$ |
| BPRS | 26.0 (3.1) | 30.8 (6.8) | 42.2 (10.9) | $\chi^2_{(2)} = 66.1, p = 4 \times 10^{-15}$ |
| SPQ | 8.6 (8.4) | 14.9 (13.3) | 30.9 (14.7) | $\chi^2_{(2)} = 54.8, p = 1 \times 10^{-12}$ |

Inclusion and exclusion criteria for this study have been reported previously (Demro et al., 2021; Schallmo et al., 2023). Briefly, PwPP included people with a confirmed diagnosis of psychotic spectrum disorders. Research diagnoses included schizophrenia (SZ, n = 35), schizoaffective disorder (SCA, n = 9), and bipolar disorder (BP, n = 20), which were assessed with the Structured Clinical Interview from the DSM-IV-TR (First, 2015). Each diagnosis was made by two clinical psychologists with expertise in psychosis spectrum disorders. For demographic information from each diagnostic group, see Supplemental Table 2. Participants in the relative group had a first degree biological relative (child, sibling, or parent) with a history of a psychosis spectrum illness but were not necessarily related to an enrolled participant in the PwPP group. Healthy control participants had no personal or immediate family history of psychosis spectrum disorders. None of our participants were adopted.

#### Exclusion Criteria

Participants were excluded if they had poorer than 20/40 visual acuity (measured with Snellen Acuity Measurement Chart (Snellen, 1862)) or other vision/hearing impairments, current or past nervous system diseases, a history of tardive dyskinesia, any condition that would inhibit task performance (e.g., paralysis or severe arthritis), electroconvulsive therapy within the last year, a head injury, skull fracture or experienced loss of consciousness greater than 30 minutes, did not speak English as a first language, could not provide informed consent, had a legal guardian, had a diagnosed learning disability or IQ less than 70, had alcohol or drug abuse within the last 2 weeks or dependence within the last 6 months, used recreational drugs or consumed more than 2 alcoholic beverages during the 24 hours leading up to the scan. Additionally, potential participants were excluded for the following MRI scanning criteria: If they had a condition that made it difficult to lie still in an MRI scanner, claustrophobia, implanted devices such as pacemakers, incontinence, head circumference larger than 62 cm, inability to fit comfortably inside the scanner (60 cm diameter bore), weight greater than 440 lb. Prior to completing the experiments included in the current study, all participants completed a series of structural MRI and fMRI scans at 3 T (Demro et al., 2021). Participants whose head motion during these 3 T fMRI scans exceeded 0.5 mm for more than 20% of TRs were excluded from all 7 T scanning procedures, including the experiments in the current study.

#### Consent and Compensation

All participants gave informed written consent prior to participation, and participants’ ability to provide informed written consent was assessed with the University of California Brief Assessment of Capacity to Consent (UBACC; Jeste et al., 2007). Experimental protocols were approved by the University of Minnesota Institutional Review Board (IRB #1607M0981) and followed guidance based on the Declaration of Helsinki’s ethical principles for medical research involving human subjects. Participants were compensated for their time.

### 2.2 Clinical Measures

Clinical measures were carried out by trained staff members under the supervision of clinicians with expertise in psychosis spectrum disorders. All participants completed the following battery of clinical questionnaires. The Brief Psychiatric Rating Scale (BPRS, 24 item version [Ventura et al., 2000] with subscale items taken from Wilson & Sponheim, 2014) assessed psychiatric symptoms of depression, anxiety, hallucinations, and unusual behavior. To examine current psychiatric symptoms, the BPRS was collected within 30 days of the 7 T visit (typically on the same day). Other clinical measures were also completed at a separate visit (see Demro et al., 2021) prior to the 7 T scanning day, which included the Brief Assessment of Cognition in Schizophrenia (BACS) to assess cognitive ability (Keefe et al., 2004) as well as the Schizotypal Personality Questionnaire (SPQ) (Raine, 1991). On the day of 7 T scanning, we also measured visual acuity using a Snellen eye chart (Snellen, 1862) (viewing distance = 100cm). Visual acuity testing was performed with corrected acuity; participants needing visual correction wore their contacts or were fitted with MRI-safe frames and lenses prior to testing.

A summary of clinical measures is shown in Table 1. For full details of our clinical methods, please refer to (Demro et al., 2021).

### 2.3 Behavioral Psychophysics

#### Equipment

Stimuli were presented in a dark room using an Apple iMac desktop and an Eizo FlexScan SX2462W monitor with a 60 Hz refresh rate (mean luminance = 61.2 cd/m^2^). A Bits# stimulus processor (Cambridge Research Systems, Kent, UK) enabled 9.6 bits of luminance resolution. Display luminance was linearized using a PR655 spectrophotometer. Participants used a height-adjustable chair and chin rest to stabilize head position at a viewing distance of 70 cm. Stimuli were generated and presented using PsychoPy v1.85.2 (Pierce, 2007).

#### Stimuli

Target stimuli were circular sine-wave luminance modulated gratings (1.1 cycles / °, 2° of visual angle in diameter, centered at 3° eccentricity along the horizontal meridian, 800 ms duration). The stimulus size of 2° of visual angle was chosen to approximate the average classical receptive field size for neurons in primary visual cortex with receptive fields centered at 3° based on studies in non-human primates (Cavanaugh et al., 2002a, 2002b; Shushruth et al., 2009, 2013) and humans (Self et al., 2016; Winawer et al., 2013) and to be sufficiently large to permit retinotopic localization using 7 T fMRI (Schallmo et al., 2016; Schumacher & Olman, 2010). Target stimuli were presented to the left and right of a central fixation mark (0.2° x 0.2° white square with a 0.1° black outline) along the horizontal meridian on a mean luminance grey background. The target gratings were surrounded by a thin, black fiducial circle with a 0.05° gap from the edge of the circular grating to help participants know where the targets would appear in space. Surrounds were 100% contrast gratings parallel to the targets (inner diameter = 2.5°, outer diameter = 4°). The edges of the target and surrounding stimuli were blurred with a raised cosine function. Target and surround gratings were presented at four different orientations (horizontal, vertical, and diagonal ±45°) and reversed contrast at 4 Hz.

In the current study, we focused on two of the conditions from our contrast surround suppression (CSS) task (Schallmo et al., 2023). Target gratings were either presented on their own (“No Surround” condition) or with a surrounding parallel annular grating (“Surround”). Both Surround and No Surround conditions were presented with a pedestal contrast of 10% (Figure 1A, adapted from Schallmo et al., 2023). On each trial a contrast increment was added to one of the two targets (left or right, randomized). The value of the contrast increment was controlled by a Psi adaptive staircase (Kingdom & Prins, 2010). In Surround trials, the surround was present around both left and right targets. In addition to the Surround and No Surround conditions, our CSS task also included 6 other conditions with no surround and different pedestal contrast levels (0, 0.65, 1.25, 2.5, 5, 20%), which enabled us to measure the classic threshold versus contrast ‘dipper’ function in all participant groups (Boynton et al., 1999; Legge & Foley, 1980; Schallmo et al., 2016; Yu et al., 2003). Results from these other conditions will be reported elsewhere in the future.

**Figure 1.**
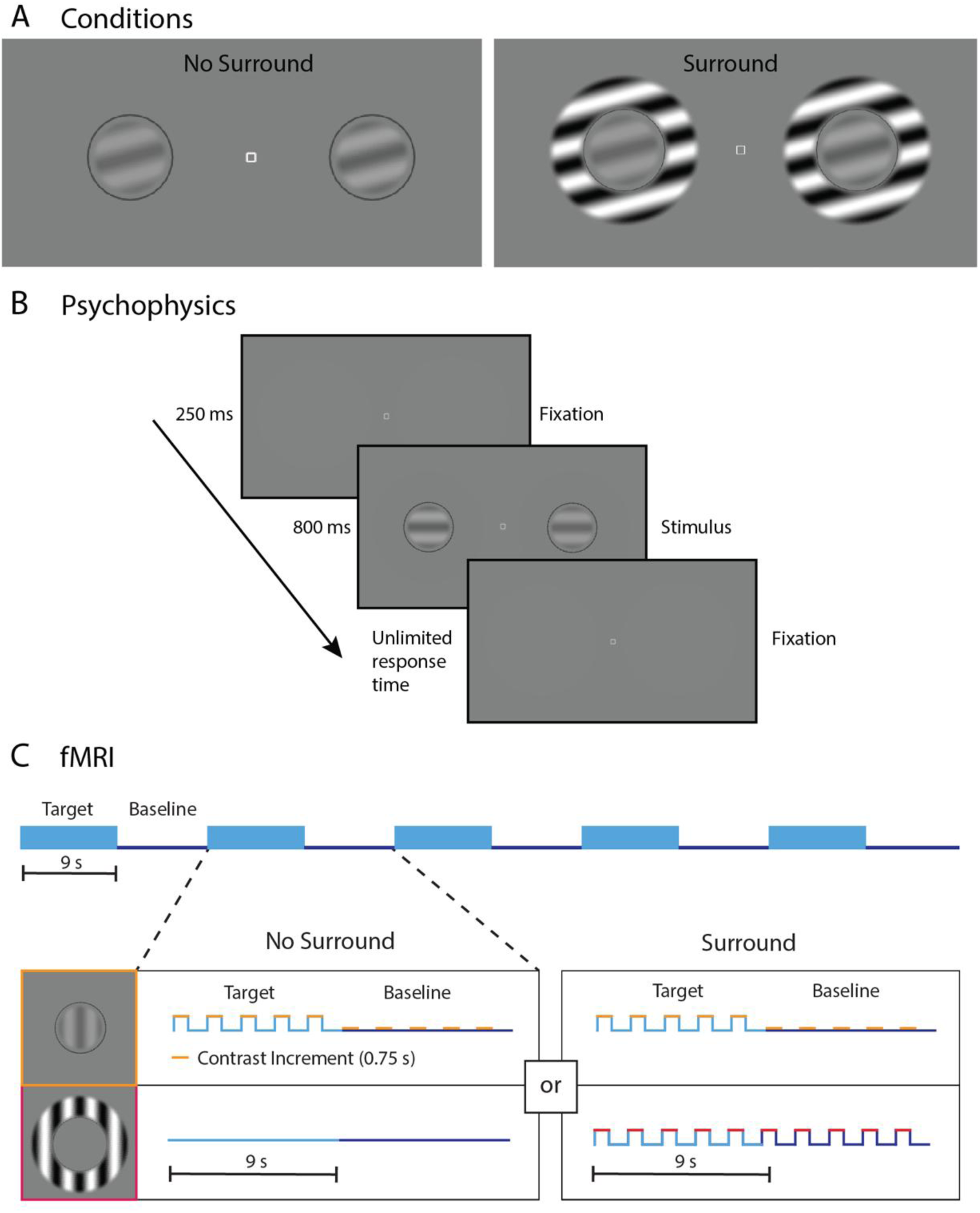
Surround suppression tasks. A) Conditions: targets were circular grating stimuli, which were either presented alone (left, No Surround) or inside surrounding parallel annular gratings to induce surround suppression (right, Surround). B) Psychophysics task: the sequence for a single trial in our psychophysical experiment. Participants were instructed to fixate on the center square and report whether the left or right grating appeared ‘stronger,’ or higher contrast. C) FMRI experimental design: we illustrate the timeline for our blocked experimental design. In both the No Surround and Surround conditions, the target (light blue) and baseline (0% contrast, dark blue) stimuli were presented in alternating 9 s blocks. Each of the 4 target contrast conditions (10, 20, 40, and 80%) was composed of five pairs of target and baseline blocks, totaling 90 s per condition (upper half of panel C). Boxes in the lower half of panel C show the timing of target (top row) and surround (bottom row) stimulus presentation within one target block (light blue) and one baseline block (dark blue), for both No Surround (left column) and Surround (right column) conditions. Each block consisted of five trials (stimulus on for 0.75 s [yellow], blank screen for 1.05 s, including the 0.6 s response period), and the participant judged which central target grating, left or right, was higher contrast. Surrounding stimuli were present in both target and baseline blocks for the Surround condition (red). Adapted from Schallmo et al., 2023.

#### Task

The CSS psychophysics task was designed to measure the effect of surrounding stimuli on psychophysical contrast discrimination thresholds and was based on stimuli from previous studies in both animal models (Angelucci & Bressloff, 2006; Cavanaugh et al., 2002b, 2002a; Shushruth et al., 2009, 2013) and humans (Schallmo et al., 2015, 2016; Zenger-Landolt & Heeger, 2003). Participants were asked to keep their eyes focused on the central fixation mark and determine which target grating (left or right) was “stronger”, or higher contrast, using their peripheral vision. They responded by pressing the left or right arrow-keys on a keyboard. In Surround trials, they were instructed to ignore the surround and make their decisions based only on the central target. If they were not certain, participants were asked to make their best guess. After reviewing the instructions and example stimuli, participants practiced the task briefly to ensure comprehension.

Each trial started with an audible tone (250 ms) and the central fixation mark, followed by the two target gratings (800 ms, with or without surrounds). The fixation mark remained on the screen until the participant responded. The response period started 400 ms after target presentation and was not limited in duration (Figure 1B). No feedback was provided.

The task was separated into eight blocks (one for each stimulus condition with pedestal contrasts at 0, 0.65, 1.25, 2.5, 5, 10, and 20% contrast with no surround, 10% contrast with surround) lasting about 4 minutes each. Each block consisted of interleaved trials from three separate staircases, which were used to estimate the contrast increment at which the participant could discriminate the location of the higher contrast grating with 80% accuracy (Kingdom & Prins, 2010; Kontsevich & Tyler, 1999). Each staircase consisted of 30 trials. Additionally, 20 catch trials—in which a fixed, high contrast increment (40%) was presented on one side—were randomly intermixed with the staircase trials in each block. Catch trials were designed to assess contrast discrimination accuracy for a low difficulty comparison, for which performance was expected to be near ceiling (e.g., to assess off-task performance or attentional lapses). Between blocks, participants were given self-timed breaks, and they were also told they could take a break during a block by withholding their response until they were ready to continue. The entire task, including instructions and practice, lasted about 40 minutes, and was performed on the same day as the fMRI experiments, prior to scanning. For one participant, psychophysical and fMRI data were acquired on separate days, due to hardware issues.

### 2.4 Functional MRI

#### Equipment

Functional MRI data were acquired using a Siemens MAGNETOM 7 T scanner equipped with an 8-kW radio frequency power amplifier and body gradients with 70 mT/m maximum amplitude and 200 T/m/s maximum slew rate (software version: VB17). Gradient echo fMRI data were collected with a Nova Medical radiofrequency 1×32 head coil (1 transmit, 32 receive channels). Our fMRI protocol had the following parameters: TR = 1 s, TE = 0.0222 s, flip angle = 45°, phase-encoding direction = anterior-posterior, resolution = 1.6 mm isotropic in 85 slices, 2-fold parallel imaging acceleration factor = 2, multiband acceleration factor = 5.

The scanner bed was fitted with head, neck, and leg support for lower back relief, and the participants were instructed to remain as still as possible during the scan. Five mm thick dielectric pads (3:1 calcium titanate powder and water) were placed under the neck and beside the temples to improve B_1_ transmit homogeneity in the cerebellum and temporal lobe regions during 7 T MRI (Vu et al., 2015).

Stimuli were generated using PsychoPy v1.85.2 (Peirce, 2007) and presented via an EPSON projector (refresh rate: 60 Hz; mean luminance: 271 cd/m^2^) on a screen mounted behind the scanner bed and viewed through a mirror mounted on the head coil (viewing distance: 100 cm). Participants responded to stimuli using two buttons on a Current Designs (Philadelphia, PA) MR-compatible 4-button response box.

For full details on our 7 T MR scanning methods, please refer to (Schallmo et al., 2023).

#### Stimuli

Stimuli for the fMRI CSS experiment were designed to measure center-surround suppression in primary visual cortex. Our fMRI paradigm mirrored that of Zenger-Landolt and Heeger (2003), but used circular target gratings with annular surrounds, rather than a concentric center-surround stimulus configuration.

Stimuli in the fMRI experiment matched those used during psychophysics (1 cycle/°, 2° of visual angle in diameter, contrast-reversing at 4 Hz, presented at 3° eccentricity from a central fixation point on a mean luminance grey background), with the following differences. There were 8 stimulus conditions of interest, plus 2 baseline conditions (10 total); target stimuli were presented at 5 contrast levels (0, 10, 20, 40, 80%) either on their own (No Surround) or with a surrounding parallel annular grating at 100% contrast (Surround). The “baseline” condition consisted of blocks with targets at 0% pedestal contrast (i.e., no target on one side and a faint reference stimulus on the other side), whereas “target on” blocks had higher pedestal contrasts. In Surround trials, both left and right targets had a surround. Grating orientation was randomized across stimulus presentations in a range of 0° to 180° in 15° increments. Stimulus duration was 0.75 s; this was 0.05 s shorter than in the psychophysics experiment to accommodate a sufficiently long response period in each trial (see below).

#### Task

The fMRI task was identical to the psychophysics task, except for the following. The response period was limited to 600 ms. Participants indicated which target they perceived to have higher contrast by pressing the left-most or right-most buttons on the button box with the thumb of the left or right hand, respectively. After a correct response, the central fixation mark turned green.

A small contrast increment was added to the target on one side, controlled by a 3-down, 1-up staircase (designed to converge at the contrast increment value the participant could discriminate with 79% accuracy (Garcıa-Pérez, 1998). This was done to ensure similar task difficulty across the fMRI and psychophysics experiments and encouraged participants to attend to the target stimuli. There was a separate contrast staircase for each of the 10 stimulus conditions (5 contrasts each for Surround and No Surround). Due to limitations of the display apparatus in the scanner, the minimum contrast increment was set to 1%. To minimize the effect of the contrast increment on fMRI response, the maximum contrast increment in each condition was set to either 25% of the condition’s pedestal contrast value or 3% contrast, whichever was greater. Because of these limitations on the stimuli, behavioral thresholds were not estimated from the participant responses gathered during the fMRI experiment.

In the fMRI task, trials were presented in a block design in cycles of target on, then target off (i.e., 0% contrast baseline). This permitted a Fourier analysis (Engel et al., 1997) to isolate the neural response to the target stimuli (Zenger-Landolt & Heeger, 2003). Trials were 1.8 s long and consisted of the stimulus being displayed for 0.75 s followed by a blank screen with a central fixation square for 1.05 s. During the blank period, participants had 0.6 s to respond. Participants were instructed that, unlike the psychophysical task, trials in the fMRI task would not wait for them to respond. Each block consisted of 5 trials of the same condition (total block duration = 9 s). For each of the 8 stimulus conditions of interest, there were 5 “target on” blocks which were interleaved with 5 “baseline” (0% contrast pedestal) blocks totaling 90 s per condition. Each condition started with a “baseline” block and ended with a “target on” block (Figure 1C). In total, there were 30 “target on” trials per condition.

Participants completed this task across 3 fMRI runs (~5 min each); each run consisted of 3 conditions. All stimulus conditions of interest (10, 20, 40, and 80% contrast, each with and without surround) was run only once during the fMRI experiment. The 3 fMRI runs were collected back-to-back in the same scanning session. The order of conditions was randomly selected from a list of 4 possible pseudo-random sequences, in which the localizer condition was always presented first.

The functional localizer was designed to identify the retinotopic regions within primary visual cortex that responded selectively to the central target stimuli and not the Surround stimulus (i.e., a differential localizer; Olman et al., 2007; Schallmo et al., 2016, 2018, 2020). This was accomplished by presenting alternating blocks of a target on, No Surround condition (target pedestal = 80% contrast, surround = 0% contrast) and a surround only condition (target pedestal = 0% contrast, surround = 100%). The trial, block, and condition timings for the functional localizer matched those in the main experiment. The functional localizer condition was contiguous with the main experiment in the first fMRI run (i.e., not a separate run).

### 2.5 Magnetic Resonance Spectroscopy

As part of the Psychosis HCP, we quantified concentrations of neural metabolites within occipital cortex during rest using magnetic resonance spectroscopy (MRS) at 7 T. A volume of interest (30 mm left-right, 18 mm anterior-posterior, 18 mm inferior-superior) was positioned in the medial occipital cortex. We measured the full MR spectrum and quantified concentrations of 18 metabolites, including the inhibitory neurotransmitter GABA. For additional details about our MRS methods, please see the Supplementary Methods section and our previous publication (Schallmo et al., 2023).

### 2.5 Data Processing and Analysis

#### Psychophysics Data Processing

Analyses were completed in MATLAB (2016a; Mathworks, Natick, MA) with the Palamedes toolbox (version 1.7.0. Prins and Kingdom, 2009). Data from the 3 staircases within each condition were pooled and fit with a Weibull function (Kingdom & Prins, 2010). The guess rate was fixed at 50%, and the lapse rate was fixed at 4%, following our previous work in healthy adults and clinical populations (Schallmo et al., 2015; Schallmo et al., 2020; Swanson et al., 2024). Analysis of catch trial accuracy indicated this value was appropriate for all participant groups (data not shown). Contrast discrimination thresholds were estimated from the fit psychometric function at 80% accuracy (Figure 2). In some cases, we found that fitting yielding unreasonable threshold estimates, which we addressed as follows. If the fit threshold was less than zero, we instead used the mean of the estimated threshold values from the last trial in each of the three staircases from that condition, or the minimum contrast value for our display (0.13%), whichever was larger. If the fit threshold was larger than the maximum contrast increment (40%) or not a real number, data from that condition were excluded.

**Figure 2.**
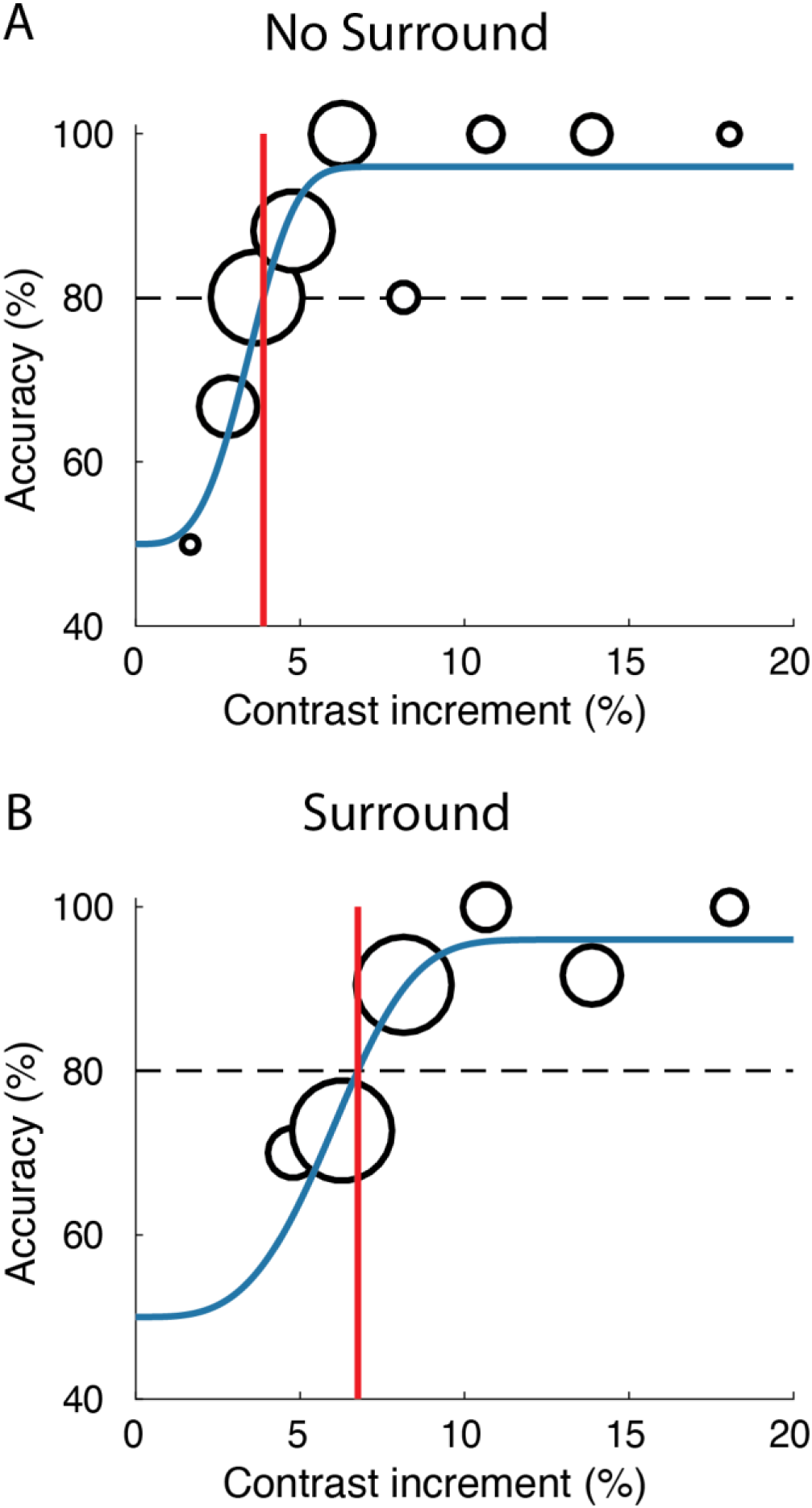
Example psychometric functions. Data from a single participant in both No Surround (A) and Surround (B) conditions. Accuracy is plotted on the y-axis and the contrast increment is plotted on the x-axis. Larger circles indicate more trials for a given contrast increment. Data were fitted with a Weibull function (blue). Thresholds (red) were estimated at 80% accuracy (dashed).

Participants with less than 80% accuracy on catch trials from all 8 psychophysical stimulus conditions were excluded from further analyses. These exclusion criteria were established *a priori*. Psychophysics data from 5 participants (all PwPP) were excluded based on these criteria. After these exclusions, catch trial accuracy did not differ significantly across groups (Kruskal Wallis, *X^2^_2_* = 1.44*, p* = 0.5). We also examined excluding participants with less than 80% accuracy in either of the 10% contrast conditions (i.e., Surround and No Surround). Excluding these individuals (1 Ctrl, 6 PwPP) did not qualitatively affect our results, thus we chose to retain their data in the main analysis. We calculated a behavioral suppression metric using the following equation:

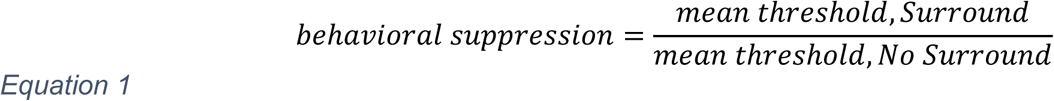

Larger values indicate stronger surround suppression, whereas values less than 1 indicate enhancement of contrast discrimination (rather than suppression). To determine whether using a ratio suppression metric would distort our pattern of results, we assessed whether the following criteria were met: 1) a linear relationship between variables in the numerator and denominator, which 2) passed through the origin (Allison et al., 1995; Curran-Everett, 2013; Mannion et al., 2017). We found that both conditions were satisfied in our data, justifying the use of a ratio suppression metric. For group comparisons, we log-transformed the suppression metrics to ensure normality. In graphs, the non-log-transformed values are plotted on a logarithmic axis.

#### FMRI Data Processing

We processed our fMRI data in AFNI version 18.2.04 (Cox, 1993), as described previously (Schallmo et al., 2023). Briefly, we used AFNI’s *3dToutcount* to identify temporal outliers and then applied slice time correction using AFNI’s *3dTshift*. We performed four volumetric preprocessing steps to correct for: 1) static gradient nonlinearities with *gradunwarp* (version 1.0.3; github.com/Washington-University/gradunwarp), 2) head motion using AFNI’s *3dvolreg*, 3) geometric distortions due to B_0_ inhomogeneity using AFNI’s *3dQwarp*, as measured in a pair of sequentially acquired gradient echo EPI scans with opposite phase encoding directions (Schallmo et al., 2021), 4) co-registration of our 7 T fMRI data to a T_1_-weighted anatomical scan (collected separately at 3 T) using AFNI’s *align_epi_anat.py*. T_1_ weighted scans were processed with FreeSurfer (version 5.3). We applied spatial smoothing (2 mm full-width half-max) using AFNI’s *3dmerge*. We performed a Fourier analysis (Engel et al., 1997) of the fMRI time series data using MATLAB and Python scripts. We used a fast Fourier transform to quantify the signal change at the block presentation frequency of 5 cycles / 90 s (0.056 Hz).

We defined regions of interest (ROIs) in the primary visual cortex (V1), the lateral geniculate nucleus of the thalamus (LGN), and the lateral occipital complex (LOC). V1 ROIs were defined from the functional localizer data (first 90 s of the first scan) using results from the Fourier analysis to identify voxels that responded more strongly to the presentation of the center target versus the surround (Engel et al., 1997; Schallmo et al., 2016). Coherence (like an un-signed correlation value) between the localizer time series data and the cyclical stimulus presentation sequence was calculated as the Fourier amplitude at 5 cycles / 90 s divided by the square root of the time series power. Voxels that were selective for the center stimulus were defined using a coherence threshold ≥ 0.2 (i.e., uncorrected *p* < 5 × 10^−5^) and phase values (which are circular between 0 - 2π) between 2.75 to 4.32, indicating that the response was in phase with stimulus presentation, after adjusting for the delayed hemodynamic response. Coherence values were chosen based on previous results, and visual inspection of early data sets in controls. We estimated the spatial profile of the noise in our fMRI data via Monte Carlo simulations using AFNI’s *3dClustSim* (Cox et al., 2017). V1 ROIs were defined in AFNI with a whole-brain cluster-corrected threshold of *p* < 0.01. V1 clusters were selected from both hemispheres in each individual data set within a V1 anatomical mask (Wang et al., 2015). Voxels in left and right hemispheres were pooled within data sets for analysis. In cases where a significant V1 cluster could be identified only in one hemisphere, data from the other hemisphere were excluded. In a small number of cases, V1 ROIs were defined by combining 2 separate clusters from within the same hemisphere based on visual inspection. Not all ROIs could be identified in all data sets. See Supplemental Table 1 for the number of data sets in which each ROI was identified.

We defined bilateral LGN ROIs using a 2 mm sphere positioned based on anatomical landmarks following visual inspection of the T_1_ weighted anatomical scan. We defined bilateral LOC ROIs based on activation during the functional localizer condition from a separate fMRI experiment focused on contour object perception task acquired within the same scanning session; for full details see Schallmo et al., (2023). LOC ROIs were identified based on the fMRI response to the presentation of an egg-shaped contour composed of Gabor elements (vs. blank) in a separate scan during the scanning session, as well as anatomical landmarks in the lateral occipital lobe (Kamath et al., 2024; Schallmo et al., 2023). LOC ROIs were defined using a Fourier analysis as above, with a coherence threshold ≥ 0.3 and a cluster corrected *p < 0.01* (Eklund 2016; Cox 2017).

#### Univariate fMRI Analysis

Further analyses were performed in MATLAB (version 2016a). We defined the following exclusion criteria for fMRI data sets: 1) excessive head motion (> 0.5 mm framewise displacement across > 20% of TR pairs across all 3 scanning runs), and 2) poor task engagement (not responding for > 10% of fMRI task trials across all 3 scanning runs). As a result, 10 data sets were excluded (1 control, 3 relatives, 6 PwPP).

For both the No Surround and the Surround conditions, we calculated the mean fMRI response within the ROI for every data set across all pedestal contrasts. Next, we quantified an fMRI suppression metric:

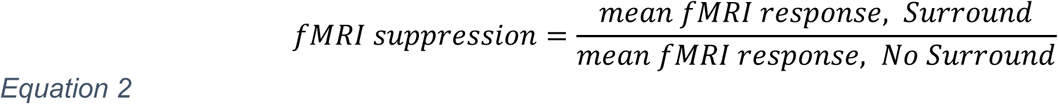

The ratio of fMRI responses in the Surround condition to the No Surround condition was log transformed to improve the normality of the distribution. Values less than 1 indicate stronger surround suppression, whereas values greater than 1 indicate enhancement. Note the opposite directionality between behavior and fMRI because we expect an inverse relationship between discrimination thresholds and fMRI responses. As for our psychophysical suppression metric, our fMRI suppression metric satisfied the criteria for undistorted ratio values (Allison et al., 1995; Curran-Everett, 2013; Mannion et al., 2017).

#### Multivariate fMRI Analyses (MVPA)

To investigate the patterns of activity across voxels in our fMRI data, we applied multivariate pattern analyses based on publicly available code (Bae & Luck, 2018). We began by defining time series epochs for each “target on” block (9 s) of our fMRI data within each ROI for each participant. Epochs were defined by taking the time series data from −2 to +15 s, relative to the target onset at the start of each block and subtracting the average signal from the baseline period (−2 to 0 s) from all time points within each individual voxel. Subtracting each voxel’s baseline signal ensured that decoding accuracy was at chance level prior to stimulus onset (data not shown). There were 5 epochs for each of the eight fMRI stimulus conditions of interest (4 pedestal contrasts for both No Surround and Surround).

We performed two different decoding analyses, in which we sought to decode the stimulus condition in a given epoch based on the fMRI time series data. In the first, we predicted the condition for each time point separately. In the second, we predicted the condition based on the average time series data from 7-9 s after stimulus onset (the time window of the peak V1 fMRI response averaged across all participants). For both analyses, we implemented support vector machine (SVM) decoding with 5-fold cross-validation using Matlab’s *fitceoc*. The SVM classifier was trained on 80% of epochs with condition labels provided, then tested on the stimulus conditions for the remaining 20% of the epochs. This was repeated 5 times, for each combination of test and training data. We performed our decoding analyses using both the real condition labels, as well as with randomly shuffled labels. The maximum number of test and training set combinations was set to 100, to limit computation time. Decoding accuracy was calculated as the percent of correctly predicted condition labels. We determined whether the decoding accuracy was significantly different from chance across in each data set by comparing the decoding accuracies we obtained from our main analyses to a null distribution generated in the shuffled decoding analysis. Data from 2 PwPP with V1 ROIs containing fewer than 10 voxels were excluded from the V1 decoding analysis.

#### Statistics

We assessed whether data were normally distributed through visual inspection, and homogeneity of variance using Levene’s test. In cases where these assumptions were met, we used repeated measures analyses of variance (rmANOVAs). Otherwise, we used Kruskal-Wallis non-parametric ANOVAs and Wilcoxon signed-rank tests. In these cases, repeated scan data from participants who returned for a second scanning session were excluded, as these tests assume independent data points. The average age and proportion of females tended to be higher among relatives compared to the other groups (Table 1). To address this, we performed post-hoc linear mixed effects models with age and sex included as co-variates in cases where we saw significant group differences with relatives. To assess the reliability of our psychophysical threshold estimates, we used two-way random effects intraclass correlation coefficients (also called ICC (3,k); Koo & Li, 2016). We investigated relationships between symptom measures and surround suppression using Spearman’s rank correlations. We quantified correlations between suppression measures (behavioral and fMRI suppression metrics, and peak decoding accuracy [7-9 s after block onset]), and BPRS, BACS, SPQ, and SGI scores. Correlation *p-*values were corrected for multiple comparisons using false discovery rate (FDR) correction. To assess whether decoding accuracy was significantly above chance across all participants, decoding results were compared to accuracy with shuffled data using paired *t*-tests.

## 3 Results

### 3.1 Behavioral Psychophysics

To assess surround suppression, we measured visual contrast discrimination thresholds (at 80% accuracy) for circular target gratings (10% contrast) presented alone or within 100% contrast surrounding annular gratings (Figure 1) in a group of healthy controls (n = 38), relatives of PwPP (n = 44), and PwPP (n = 64). As expected, we observed a strong surround suppression effect (ANOVA, *F*_1,134_ = 250, *p* < 0.001); thresholds were higher on average with the Surround vs. No Surround (mean: 10.6% vs. 3.6%; Figure 3), indicating that participants needed a larger contrast difference to reliably discriminate targets with surrounds. There was also a significant group difference in thresholds overall (*F*_2,134_ = 4.98, *p* = 0.008), such that PwPP had higher thresholds on average compared to relatives and controls, regardless of the presence of the surround (post hoc ANOVA, control vs. PwPP: *F*_1,91_ = 7.3, *p* = 0.008; relatives vs. PwPP: *F*_1,102_ = 4.0, *p* = 0.047). However, we found that the difference in thresholds between PwPP and relatives was no longer significant after including sex and age as co-variates (linear mixed effects model, *t*_257_ = 0.64, parameter estimate [SE] = 0.03 [0.053], *p* = 0.5). We found no significant interactions between group and surround (across all participants: *F*_2,124_ = 1.04, *p* = 0.4; PwPP vs. HC only: *F*_1,84_ = 1.59, *p* = 0.2), contrary to our prediction of weaker surround suppression among PwPP. Threshold estimates showed high test-retest reliability across 3 independent measurements (i.e., staircases) in both Surround and No Surround conditions (ICC = 0.82 and 0.83 respectively, Supplemental Figure 1A & B). Across experimental sessions separated by several months, threshold estimates in PwPP were fairly stable for the Surround condition (ICC = 0.74), but longitudinal variability was higher for the No Surround condition (ICC = 0.43, Supplemental Figure 1C & D).

**Figure 3.**
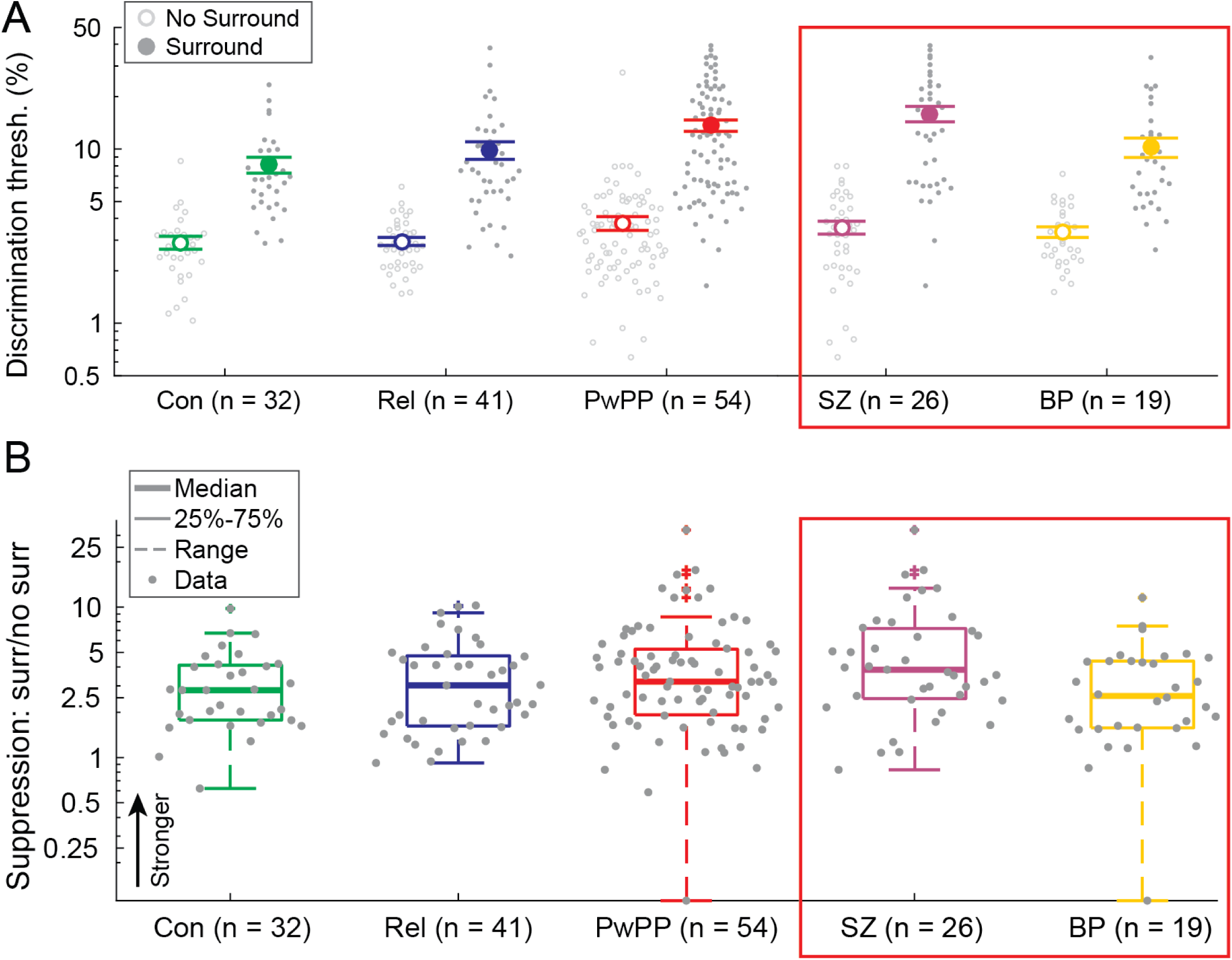
Psychophysical results. A) Contrast discrimination thresholds (No Surround: open symbols, Surround: closed) are shown for controls (Con; green, n = 32), relatives (Rel; blue, n = 41), and PwPP (red, n = 54) on a log axis. In the red box, we show a breakdown of data from individuals in the PwPP group with schizophrenia (SZ; purple, n = 26) and bipolar disorder (BP; yellow, n = 19). Psychophysical data from repeat sessions are included for 29 PwPP (13 SZ, 13 BP). To better illustrate the main results, data from 1 participant with BP are not shown (i.e., outside the y-axis limits). B) Suppression metrics (Surround / No Surround) for each group on a log axis. Thick lines show the median, box edges show 25 and 75%, dashed lines show 1.5x the interquartile range, gray dots show individual data points. Larger values indicate stronger suppression (upward arrow).

To further examine surround suppression in our contrast discrimination task, we calculated a normalized suppression metric for each data set as the ratio of mean thresholds in the Surround condition vs. mean thresholds for No Surround (Equation 1). We saw no significant group differences in suppression metrics (Kruskal-Wallis, *X*^2^ = 2.57, *p* = 0.3, Figure 3), which again was contrary to our prediction of weaker suppression among PwPP. We also did not find any significant associations between surround suppression metrics and BPRS, BACS, SPQ, or SGI scores (Spearman’s correlations across all participants, |*r*| values < 0.11, uncorrected *p* values > 0.2; across PwPP only, |*r*| values < 0.24, uncorrected *p* values > 0.10). Thus, results from our behavioral surround suppression task did not show the expected effect of weaker suppression among PwPP, nor any significant associations with measures of psychotic psychopathology.

We next asked whether diagnostic sub-groups among PwPP (i.e., schizophrenia, SZ, n = 35; and bipolar disorder, BP, n = 20) might show differences in contrast surround suppression. We excluded participants with schizoaffective disorder (n = 9) from this analysis, due to the small sample size. We observed a significant difference in contrast discrimination thresholds between groups (ANOVA, *F*_2,81_ = 4.39, *p* = 0.015) such that thresholds were significantly higher in the SZ group vs. controls (post hoc ANOVA, *F*_1,62_ = 10.2, *p_uncorr._* = 0.002, *p_FDR_* = 0.006). In this case, we saw a significant interaction between group and surround for threshold values (ANOVA, *F*_2,81_ = 3.55, *p* = 0.034), and suppression metrics also differed significantly between controls, SZ, and BP participants (Kruskal-Wallis, *X*^2^_2_ = 6.55, *p* = 0.038, Figure 3). Post hoc tests revealed suppression was greater in the SZ vs. BP group (Wilcoxon; *Z* = −2.29, *p_uncorr._* = 0.022, *p_FDR_* = 0.044). There was also a trend towards greater suppression in the SZ group vs. controls (*Z* = −1.96, *p_uncorr._* = 0.019, *p_FDR_* = 0.056). Stronger suppression in SZ vs. controls remained significant with age and sex included as co-variates (linear mixed effects model, *t*_210_ = 2.42, parameter estimate [SE] = 0.19 [0.078], *p* = 0.016), whereas the difference in thresholds between SZ vs. controls did not (*t*_210_ = 0.87, parameter estimate [SE] = 0.06 [0.065], *p* = 0.4). These results suggest that in our psychophysical task, surround suppression was stronger among participants with SZ versus BP and controls, contrary to our prediction of weaker suppression among PwPP.

### 3.2 Functional MRI

#### Univariate Analysis

To investigate neural surround suppression, we measured 7 T fMRI responses among controls (n = 37), relatives (n = 36), and PwPP (n = 49) during a contrast discrimination task similar to our psychophysical paradigm (Figure 1). Here, we used 8 stimulus conditions: 10, 20, 40, and 80% contrast targets, each with and without a surrounding annulus (100% contrast). Stimuli were presented in a cyclical block design that allowed us to isolate and quantify the fMRI response to target gratings (Zenger-Landolt & Heeger, 2003). We examined fMRI responses in three ROIs: V1, LGN, and LOC. These ROIs were chosen based on their known roles in the visual processing hierarchy, and because previous studies have suggested they may play a role in visual perceptual dysfunction among PwPP (Anderson et al., 2017; Seymour et al., 2013; Silverstein et al., 2015).

As expected, higher contrast stimuli yielded higher V1 fMRI responses (ANOVA, *F*_1,118_ = 126.5, *p* = 3 × 10^−21^, Figure 4) and the surround attenuated V1 fMRI responses across contrasts (*F*_1,118_ = 38.2, *p* = 7 × 10^−9^, Figure 4). V1 fMRI responses (across all conditions) differed significantly across groups (*F*_2,118_ = 7.15, *p* = 0.001), with overall higher fMRI responses for PwPP versus controls (post-hoc repeated measures ANOVA, *F*_1,68_ = 5.76, *p_uncorr_* = 0.018, *p_FDR_* = 0.055). There was also a significant interaction between surround and contrast (*F*_1,119_ = 9.49, *p* = 0.003) such that surround suppression was stronger at higher contrasts across all 3 groups. Other interactions were not significant.

**Figure 4.**
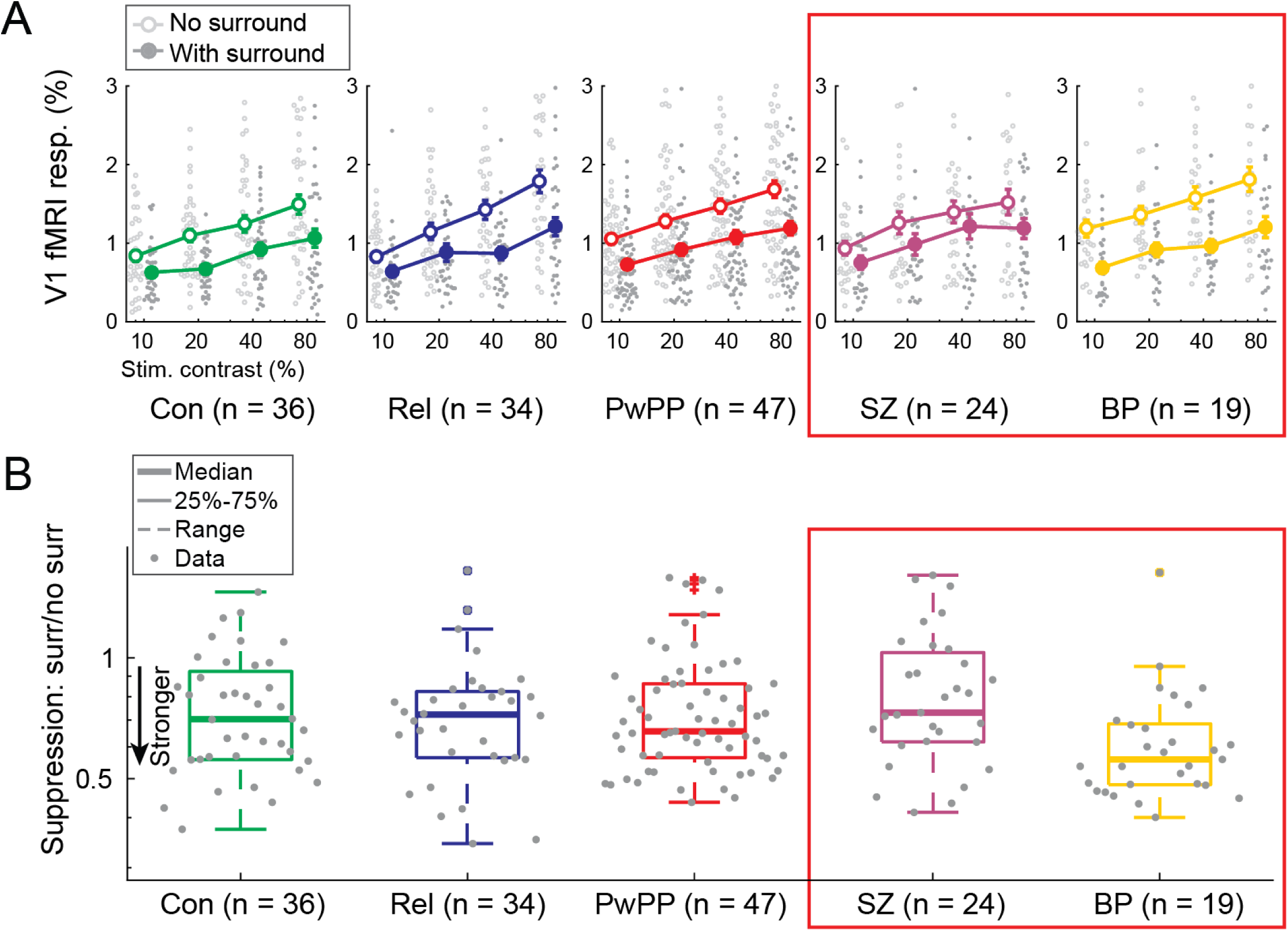
V1 fMRI results. A) FMRI responses in V1 from controls (Con; green, n = 36), relatives (Rel; blue, n = 34), and PwPP (red, n = 47) for No Surround (open) and Surround (closed) conditions at four pedestal contrast levels (10%, 20%, 40%, 80%; x axis). Light (No Surround) and dark grey (Surround) dots show individual data points, larger circles show group means. Error bars are standard error of the mean. Red box shows a breakdown of data from PwPP with schizophrenia (SZ; purple, n = 24) and bipolar disorder (BP; yellow, n = 19). Data from repeat scanning sessions are included for 4 controls and 20 PwPP (7 SZ, 10 BP, 3 SCA). B) V1 fMRI suppression metrics (Surround / No Surround; smaller values indicate stronger suppression). Thick lines show the median, box edges show 25 and 75%, dashed lines show 1.5x the interquartile range, gray dots show individual data points.

To quantify the magnitude of surround suppression in the fMRI response, we calculated a suppression metric for each data set (Equation 2). Similar to our behavioral experiment, we expected V1 suppression to be attenuated among PwPP versus controls, but we found no significant group differences in V1 fMRI suppression metrics (ANOVA, *F*_2,119_ = 0.254, *p* = 0.8, Figure 4B). However, when dividing PwPP into diagnostic sub-groups, there was a significant difference in suppression between groups (*F*_2,76_ = 3.90, *p* = 0.024) driven by higher fMRI suppression in the BP group compared to SZ (post hoc ANOVA, *F*_1,41_ = 7.95, uncorrected *p* = 0.007, FDR corrected *p* = 0.021). Thus, we again found differences in suppression between our diagnostic groups, which differed from our prediction of weaker surround suppression among PwPP versus controls. In this case, suppression of the V1 fMRI response showed the opposite pattern (stronger for BP vs. SZ) as compared to our psychophysical results (stronger for SZ vs. BP). We did not observe the expected effect of significantly weaker suppression in SZ vs. controls.

FMRI responses in the LGN (across all conditions) were higher among PwPP and relatives than controls (ANOVA, *F*_2,118_ = 10.3, *p* = 7 × 10^−5^; post hoc ANOVA PwPP vs. controls, *F*_1,83_ = 17.2, *p_uncorr._* = 7 × 10^−5^, *p_FDR_ =* 2 × 10^−4^; post hoc ANOVA relatives vs. controls, *F*_1,70_ = 5.2, *p_uncorr_*. = 0.026, *p_FDR_ =* 0.052). Response magnitude increased slightly with stimulus contrast (*F*_1,118_ = 4.8, *p* = 0.030, Supplemental Figure 2). However, there was no significant surround suppression (*F*_1,118_ = 0.004, *p* = 1.0) in the LGN, and no significant interactions. Given the lack of a main effect of surround, we did not examine fMRI suppression metrics from the LGN.

As expected, LOC fMRI responses were lower in Surround vs. No Surround conditions (ANOVA, *F*_1,104_ = 34.3, *p* = 4 × 10^−8^, Figure 5) and LOC fMRI responses increased with higher stimulus contrast (*F*_1,104_ = 16.5, *p* = 9 × 10^−5^). Overall, LOC fMRI response amplitudes did not differ significantly across groups (3-way ANOVA, F_2,104_ = 2.1, *p* = 0.13). There was a significant interaction between surround and contrast (*F*_1,104_ = 9.3, *p* = 0.003) with greater suppression at higher contrasts. No other interactions were significant. Of note, suppression metrics in LOC were weaker in both PwPP and relatives compared to controls (ANOVA, *F*_2,104_ = 4.60, *p* = 0.012, Figure 5; post hoc ANOVA, control vs. PwPP, *F*_1,69_ = 9.7, *p_uncorr._* = 0.0025, *p_FDR_* = 0.0075; control vs. relative, *F*_1,63_ = 6.64, *p_uncorr._* = 0.012, *p_FDR_* = 0.025). Post hoc linear mixed effects analyses showed that differences in LOC suppression were not explained by group differences in age or sex (linear mixed-effects model, *t*_64_ = 3.11, parameter estimate [SE] = 0.14 [0.047], *p* = 0.003). Further, LOC suppression was weaker in participants with SZ compared to controls (post hoc ANOVA, controls vs. SZ, *F*_1,46_ = 12.4, *p_uncorr._* = 9 × 10^−4^, *p_FDR_* = 0.0029) whereas BP participants showed intermediate suppression (post hoc ANOVA, controls vs. BP, *F*_1,46_ = 3.48, *p_uncorr_*. = 0.056, *p_FDR_* = 0.17). Weaker suppression in LOC correlated with higher psychiatric symptoms (BPRS) and poorer cognition (BACS), but these did not survive correction for multiple comparisons (see Supplemental Results for further details). Thus, we observed the expected pattern of reduced surround suppression in PwPP compared to controls for fMRI responses in area LOC, but not in V1 or the LGN.

**Figure 5.**
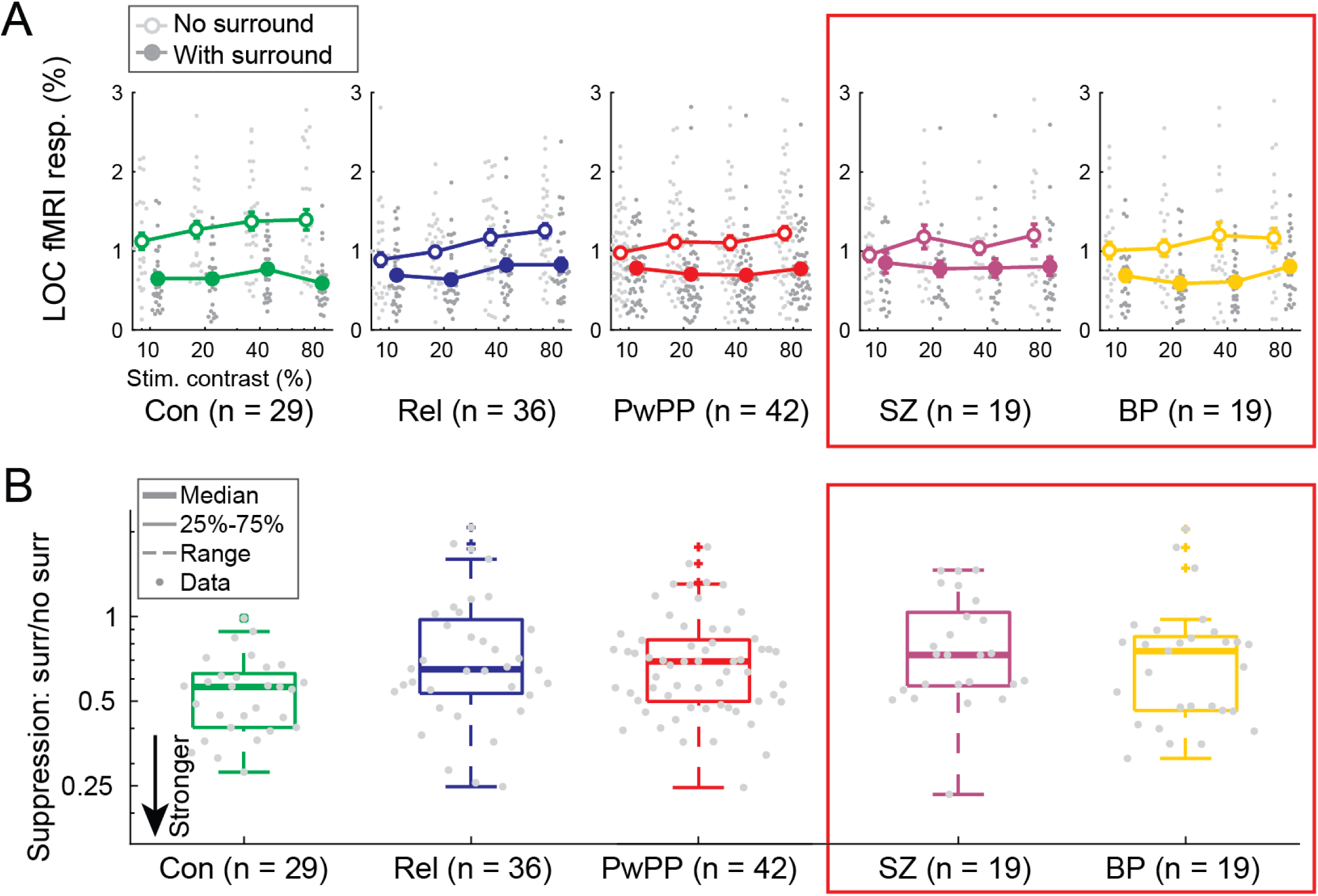
LOC fMRI results. A) FMRI responses in LOC from controls (Con; green, n = 29), relatives (Rel; blue, n = 36), and PwPP (red, n = 42) for No Surround (open) and Surround (closed) conditions at four pedestal contrasts (10%, 20%, 40%, 80%; x axis). Light (No Surround) and dark grey (Surround) dots show individual data points, larger circles show group means. Error bars are standard error of the mean. Red box shows a breakdown of data from PwPP with schizophrenia (SZ; purple, n = 19) and bipolar disorder (BP; yellow, n = 19). Data from repeat scanning sessions are included for 19 PwPP (6 SZ, 10 BP, 2 SCA). B) LOC fMRI suppression metrics (Surround / No Surround; smaller values indicate stronger suppression). Thick lines show the median, box edges show 25 and 75%, dashed lines show 1.5x the interquartile range, gray dots show individual data points.

#### Multivariate Analysis

Although we found no significant group differences in the magnitude of fMRI suppression within V1, multivariate analyses are sensitive to spatial heterogeneity of responses within individuals that can be missed by univariate analyses (Davis et al., 2014; Haynes, 2015). Whereas univariate analyses are sensitive to group differences in activation level averaged across voxels within the ROI, multivoxel pattern analysis captures the information contained within patterns of voxel-wise activity within subjects. Thus, we examined whether multi-voxel patterns of activity within V1 contained reliable information that could differentiate the stimulus conditions. Our multivoxel pattern analyses used support vector machine decoding with 5-fold cross validation (i.e., training on data from 4 of 5 fMRI blocks and testing on the 5^th^). Here, we focus on our results when decoding the presence or absence of a surrounding stimulus based on the fMRI response to the target in the center (for additional information about the results of our decoding analyses, please see Supplemental Results). Across all participants, accuracy for decoding Surround vs. No Surround (collapsing across contrasts) during the peak V1 fMRI response (i.e., 7-9 s after block onset) was significantly greater than chance (vs. decoding with shuffled labels; paired *t*-test, *t*_141_ = −6.88, *p* < 0.0001). Importantly, decoding accuracy for Surround vs. No Surround from V1 fMRI responses differed between groups (ANOVA, *F*_2,139_ = 3.62, *p* = 0.029, Figure 6), which appeared driven in part by lower accuracy in the PwPP group vs. controls, though this post-hoc comparison did not reach statistical significance (ANOVA, *F*_1,110_ = 3.59, *p_uncorr._* = 0.061). Poor Surround vs. No Surround decoding among PwPP was not explained by a difference in the univariate fMRI response, which was comparable across groups (Supplemental Figure 4). Instead, our results may reflect subtle differences in the pattern of activity across voxels with versus without surrounding stimuli within the region of V1 that responded to the target gratings.

**Figure 6.**
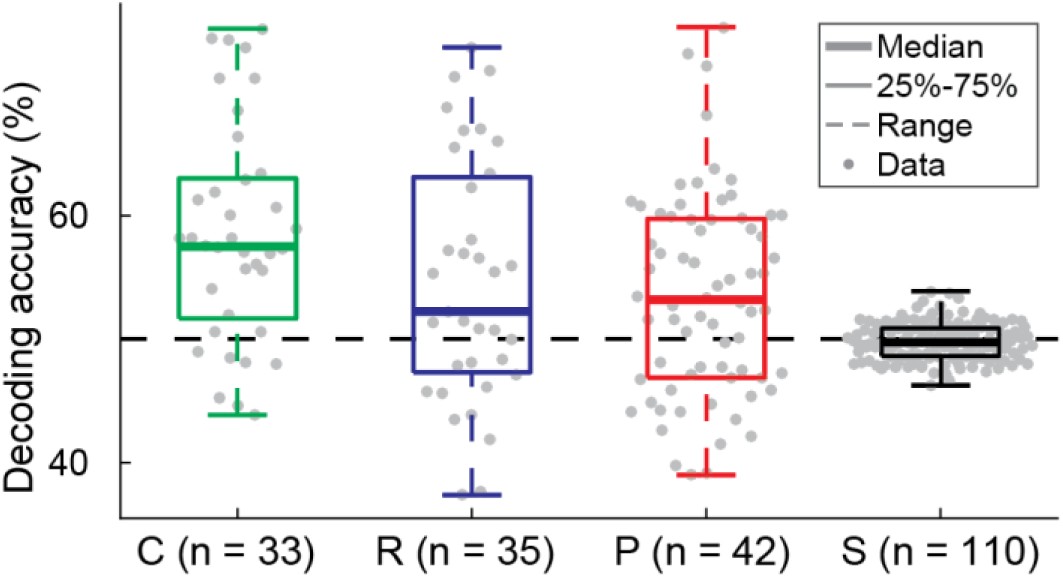
V1 fMRI decoding accuracy for Surround vs. No Surround. Decoding accuracy for average fMRI response 7-9 s post stimulus onset in each group (Control [C], n = 33; Relatives [R], n = 35; and PwPP [P], n = 42, plus repeated scans from 4 Controls and 20 PwPP). Boxplots show median (thick line), 25%, and 75% quartiles (box). Whiskers show 1.5x interquartile range. Gray dots show individual data points. The dashed line shows chance decoding accuracy (50%), and the shuffled (S, n = 110) data shows decoding accuracy for a classifier trained on data with shuffled labels.

An examination of associations between V1 surround decoding accuracy and measures of psychiatric symptoms revealed a correlation between higher BPRS scores and poorer V1 surround decoding that did not survive correction for multiple comparisons (see Supplemental Results for further details). We saw no significant group differences in decoding of fMRI responses from the LGN or LOC (see Supplemental Results).

#### Magnetic Resonance Spectroscopy

Finally, we examined whether the magnitude of surround suppression was related to individual or group differences in the concentration of GABA, an inhibitory neurotransmitter, within visual cortex, as measured by MR spectroscopy (MRS). Previous work has suggested that weaker surround suppression may be associated with lower levels of the inhibitory neurotransmitter GABA, including among people with schizophrenia (Cook et al., 2016; Yoon et al., 2010) but see (Schallmo et al., 2018; Schallmo et al., 2020) for conflicting results. We saw no significant group differences in occipital GABA levels at rest between controls, relatives, and PwPP (*Χ*^2^_2_ = 0.09 *p* = 1.0; Supplemental Figure 8A). There were also no significant correlations between GABA levels and suppression indices from our behavioral task across participants from all groups (Spearman’s correlation, *r*_117_ = −0.003, *p* = 1.0; Supplemental Figure 8B) nor within each group individually (|*r* values| < 0.26, *p_uncorr_*. values > 0.07, *p_FDR_* values > 0.2). As such, we did not find evidence to support a direct relationship between GABA levels and the magnitude of surround suppression in our task. Similarly, we found no evidence for significant correlations between GABA levels and suppression indices from V1 fMRI across all participants (Spearman’s correlation, *r*_108_ = −0.03, *p* = 0.8; Supplemental Figure 8C) nor within each group individually (|*r* values| < 0.25, *p_uncorr_*. values > 0.15, *p_FDR_* values > 0.4).

## 4 Discussion

We investigated whether surround suppression differed between PwPP, first degree biological relatives, and healthy controls using both psychophysics and 7 T fMRI in the LGN, V1, and LOC. Across participants, we saw a robust behavioral surround suppression effect: higher contrast discrimination thresholds for targets with surrounds vs. no surround. Surprisingly, we found no group differences in the magnitude of this suppression between PwPP, relatives, and controls, contrary to our hypothesis that PwPP would show weaker suppression. However, when we divided our PwPP group by diagnosis, behavioral surround suppression was greater in participants with SZ compared to BP, with a trend toward stronger suppression for SZ vs. controls. This suggests there may be important differences in perception of surround suppression across diagnoses that involve psychosis. Similar to our behavioral results, we saw robust surround suppression of the V1 fMRI response across participants, but there were no significant group differences in the magnitude of V1 suppression. When comparing PwPP across diagnostic categories, we again found significant group differences, this time with stronger V1 fMRI suppression in the BP group compared to SZ (this effect is in the opposite direction of the behavioral result). We used a decoding analysis to investigate whether there might be discernable group differences in the multivariate pattern of V1 neural activity between Surround and No Surround conditions. We saw a significant difference across all 3 groups, with a trend toward lower accuracy in decoding Surround vs. No Surround among PwPP compared to controls, suggesting less reliable differences in the neural representation of surrounding spatial context within V1 in psychosis. Interestingly, surround suppression in area LOC was weaker in both PwPP and relatives compared with controls, which followed the expected pattern. LOC suppression was also weaker in people with SZ versus controls.

### 4.1 Relation to Prior Literature

#### Behavior

We expected to find reduced perceptual surround suppression in PwPP vs. controls, as weaker context-dependent contrast illusions have been observed across many studies in both schizophrenia (Kéri et al., 2005; Must et al., 2004; Serrano-Pedraza et al., 2014) and to a lesser extent in bipolar disorder (Schallmo et al., 2015; Yang et al., 2013). A recent meta-analysis found evidence for weaker surround suppression during contrast perception in SZ, reporting an effect size of Hedge’s *g* = 0.6 for tasks such as ours (Pokorny, Klein, et al., 2024). However, it is worth noting that there are a few studies that have found stronger effects of surrounding spatial context in SZ vs. controls, with regard to motion and orientation perception (Chen et al., 2008; Thakkar et al., 2021). One study from the Cognitive Neuroscience Test Reliability and Clinical applications for Schizophrenia (CNTRaCS) Consortium, which focused on stable outpatients and had a large sample size (n = 132 SZ, n = 130 controls), found only a small effect of reduced surround suppression during contrast perception in SZ, and this was only marginally significant after excluding participants with frequent attentional lapses (i.e., < 70% accuracy on catch trials; Barch et al., 2012). Another recent study from our group found that the difference in surround suppression between SZ and controls was moderated by visual acuity (Pokorny et al., 2023). In the current study, we recruited stable outpatients (for whom symptom severity may be lower), we excluded participants who performed poorly on catch trials (< 80% correct), and our groups were matched on visual acuity (Table 1), all of which may have limited our ability to detect a group difference in surround suppression in our behavioral task.

#### Functional MRI

We also expected to see weaker surround suppression of the V1 fMRI response in PwPP vs. controls, in line with the results of Seymour and colleagues (2013). However, we note that their study measured suppression when the surrounding stimulus appeared and disappeared across blocks, while the center stimulus remained on the screen. When measuring blood oxygen level dependent (BOLD) responses within a retinotopic target ROI, such a design may confound direct effects of neural surround suppression with indirect suppression of the BOLD response (e.g., reduced blood flow within the target ROI by activation in surrounding cortex, as a result of vascular signal displacement or blood stealing; Olman et al., 2007; Shmuel et al., 2002). We sought to address this issue in our study by measuring fMRI responses to blocks of target present vs. target absent, with surrounding stimuli held constant across blocks, and then comparing response magnitudes with and without surrounds. Another study used fMRI to quantify population receptive field sizes, and found smaller suppressive pRF surrounds in SZ (n = 13) vs. controls (n = 14) in areas V1, V2, and V4 (Anderson et al., 2017). However, a recent study using fMRI at 7 T found no significant difference in V1 surround suppression between people with schizophrenia, bipolar disorder, and controls (Pokorny, Sponheim, et al., 2024). Our fMRI results are partially consistent with these previous findings; although we did not find group differences in V1 suppression, V1 decoding accuracy trended lower, and suppression in LOC was significantly weaker in PwPP vs. controls (in the expected direction).

#### The Role of Stimulus Parameters and Task Design

The magnitude of surround suppression can vary as a result of small but important differences in stimuli and task design that may influence the participant’s ability to segregate center and surround (Lotto & Purves, 2001; Schallmo et al., 2015; Xing & Heeger, 2000; Yu et al., 2001). In the current study, we used relatively small center (2° diameter) and surrounding (1.5° wide) stimuli, presented simultaneously and oriented parallel to one another. In some cases, previous studies have found significantly weaker surround suppression in PwPP when surround orientation differed from the center (e.g., orthogonal), but group differences were smaller or absent with parallel surrounds (Pokorny et al., 2023), which might have limited our ability to detect group differences in the current study.

Effects of surround suppression may also be difficult to distinguish from general impairments in contrast sensitivity in PwPP. In our task, participants were instructed to judge which center grating (left or right) was higher contrast (i.e., discrimination), and surrounds, when present, appeared on both sides. Surrounding stimuli made perception of target contrast more difficult, yielding higher discrimination thresholds. While measuring discrimination thresholds can enable inferences about the underlying neural contrast response (Zenger-Landolt & Heeger, 2003), discrimination thresholds may be higher in PwPP even *without* surrounding stimuli (Yoon et al., 2009), as a result of generally impaired contrast sensitivity (Butler et al., 2008; Skottun & Skoyles, 2007). Thus, when examining contrast discrimination in PwPP, it may be difficult to differentiate effects of impaired contrast sensitivity in general versus surround suppression *per se* (see Mannion et al., 2017 for further consideration of this issue).

Relatedly, stimulus and task parameters may have influenced the deployment of spatial attention in our paradigm. Surround suppression depends on attention (Flevaris & Murray, 2015; Motoyoshi et al., 2015; Reynolds & Heeger, 2009), with narrower spatial attention yielding weaker suppression (Kınıklıoğlu & Boyaci, 2022; Schallmo et al., 2020). Spatial attention is generally thought to be altered in schizophrenia, but there is disagreement as to the nature of this disruption; proposals include poorer distractor filtering (Hahn et al., 2022; Luck, Leonard, et al., 2019), attentional hyperfocusing (Hahn et al., 2022; Leonard et al., 2017; Luck, Hahn, et al., 2019) and impaired attentional orienting or control (Lynn & Sponheim, 2023). For example, attentional control may be poorer when target and distractor stimuli are more similar, whereas people with schizophrenia may focus attention more narrowly when competition from distractors is weaker (Hahn et al., 2022). In our task, we would expect that narrower spatial attention would yield weaker surround suppression among PwPP, whereas impairments in attentional control, selection, or filtering of distractors would yield stronger surround suppression (e.g., spreading attention to the surround could increase the magnitude of suppression directly, or participants may struggle to base their judgments on information from the target rather than the surround during discrimination). As effects in both directions appear plausible, the overall effect of surround suppression in PwPP for a given task may depend on stimulus and task parameters that influence the allocation of spatial attention, and the ability to perceptually segment center and surrounding stimuli (Pokorny, Sponheim, et al., 2024). Future studies may elucidate the role of spatial attention in surround suppression among PwPP by explicitly manipulating deployment of attention across task conditions (e.g., Kınıklıoğlu & Boyaci, 2022).

#### Relation to GABA

Previous work has suggested that weaker surround suppression in SZ may be linked to altered inhibition via the neurotransmitter GABA (i.e., lower concentrations) in early visual cortex (Cook et al., 2016; Song et al., 2021; Yoon et al., 2010). Using MR spectroscopy at 3 T, Yoon and colleagues (2010) found lower concentrations of GABA in visual cortex among people with schizophrenia (n = 13) vs. controls (n = 13). Across a subset of participants (9 controls, 7 SZ), lower GABA levels correlated with weaker surround suppression (measured behaviorally as the ratio of contrast discrimination thresholds for targets with parallel vs. orthogonally oriented surrounds). In another study of healthy controls (n = 9), Cook and colleagues (2016) found that lower GABA levels (again measured with MRS at 3 T) correlated with weaker surround suppression (measured behaviorally as the ratio of perceived contrast for targets with surrounds vs. no surrounds). Both of these studies are limited by small sample sizes. Another limitation of the MEGA-PRESS MRS technique (Mescher et al., 1998) used in both studies is the inclusion of co-edited macromolecule signals with the GABA peak at 3 ppm (Mullins et al., 2014). We sought to avoid this limitation in the current study using an MRS approach at 7 T without spectral editing (Schallmo et al., 2023) and by including macromolecule signals in our basis set measured using inversion recovery methods (Behar et al., 1994; Marjańska et al., 2017). Unlike the previous studies mentioned above, we found no significant correlations between behavioral or V1 suppression metrics and occipital GABA levels within or across groups. Thus, we did not find any evidence in the current study to support a link between reduced occipital GABA and impaired surround suppression in PwPP.

### 4.2 Strengths and Limitations of the Current Study

The strengths of the current study include the large sample sizes (n = 64 PwPP, 38 controls), and the inclusion of biological relatives of PwPP (n = 44), which permitted us to examine the role of genetic liability for psychosis in the absence of a diagnosed psychotic illness. This is the largest neurophysiological study of center-surround processing in PwPP to date. By making our data and experimental code publicly available, our study should facilitate reproducibility. We chose a trans-diagnostic approach to studying visual perception in psychotic psychopathology. Many previous studies of surround suppression in people with psychotic disorders have separated participants according to diagnostic category, or focused on a single diagnosis (e.g., SZ). However, the diagnostic categories defined by the Diagnostic and Statistical Manual of Mental Disorders have been criticized for having relatively poor reliability and validity (Kotov et al., 2017; Markon et al., 2011). Here, we investigated visual dysfunction in psychotic psychopathology more broadly. Our approach was informed by the National Institute of Mental Health’s Research Domain Criteria (RDoC) framework (Cuthbert, 2014), and by the notion that visual dysfunction may exist on a spectrum that spans across psychotic disorders, and may be present among biological relatives of PwPP.

Using ultra-high field functional MRI and MR spectroscopy at 7 T allowed us to investigate the physiological basis of perceptual surround suppression in PwPP, relatives, and controls. Only a few studies have used brain imaging to examine surround suppression in PwPP, and even fewer have been performed at 7 T. Conducting our study at 7 T provided us with higher fidelity (i.e., BOLD contrast-to-noise, signal-to-noise for MR spectroscopy) as compared to similar approaches at 3 T (Godlewska et al., 2017; Terpstra et al., 2016; Vu et al., 2015). An additional advantage of this study is our multi-modal approach combining behavioral psychophysics, 7 T fMRI, and 7 T MRS in the same participants.

There are several limitations to consider as well. First, our relative group tended to be older and had a higher proportion of individuals assigned female at birth compared to the control and PwPP groups.

To address this, we included age and sex at birth as factors in post hoc analyses in cases where there were significant differences between relatives and other groups. Our recruitment strategy may also have introduced a selection bias, as we only recruited participants for the current experiments if they had previously completed 3 T scanning as part of the Psychosis Human Connectome Project (Demro et al., 2021), and did not show excessive head motion during the preceding fMRI scans. This criterion may have excluded more symptomatic individuals in the PwPP group, which may have limited our ability to detect group differences related to symptom severity. Additionally, while our transdiagnostic approach was one of the goals of the Psychosis Human Connectome Project, this limited the number of participants within each diagnostic category, therefore limiting our statistical power to compare between diagnostic groups within PwPP. However, we found some interesting differences in behavior and V1 fMRI suppression between our SZ and BP groups, suggesting further research comparing surround suppression between these groups may be warranted.

## Data Availability

Unprocessed imaging (i.e., DICOM) data, behavioral data, and clinical data will be available from the National Data Archive (nda.nih.gov/edit_collection.html?id=3162). Processed data will be made available by the authors upon request. Experimental task code and data processing code are available on GitHub (github.com/mpschallmo/PsychosisHCP).

https://github.com/mpschallmo/PsychosisHCP

## Acknowledgments

The authors would like to thank the following individuals for their support with data collection and / or processing: Philip C. Burton, Victoria Espensen-Sturges, Kyle W. Killebrew, Marisa J. Sanchez, and Kimberly B. Weldon. The Psychosis Human Connectome Project team also included: Jessica Arend, Haven Hafar, Isaac Hatch-Gillette, Elijah Lahud, Tim Lano, Evan Myers, Karina Smiley, Yeliz Toker, Brianna Wenande, Casey Xamonthiene, and Alina Yasis.

## 6 Supplemental Materials

### Supplemental Methods

**Supplemental Table 1:**
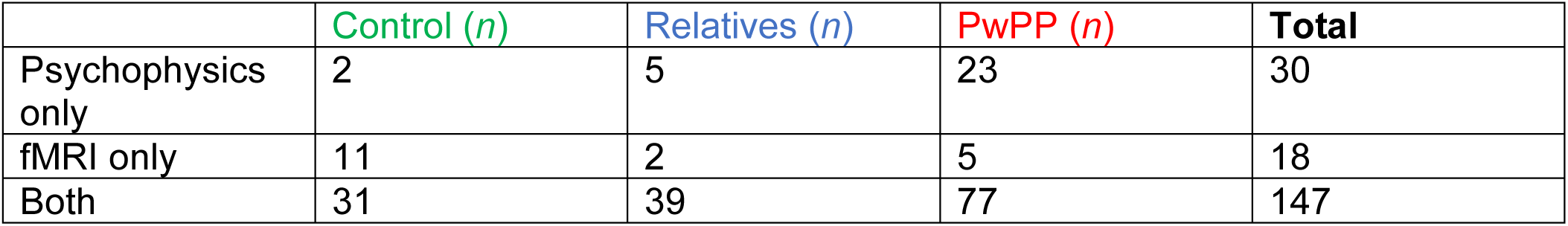
Number of datasets completed for psychophysics only, fMRI only, or both fMRI and psychophysics by group.

**Supplemental Table 2:**
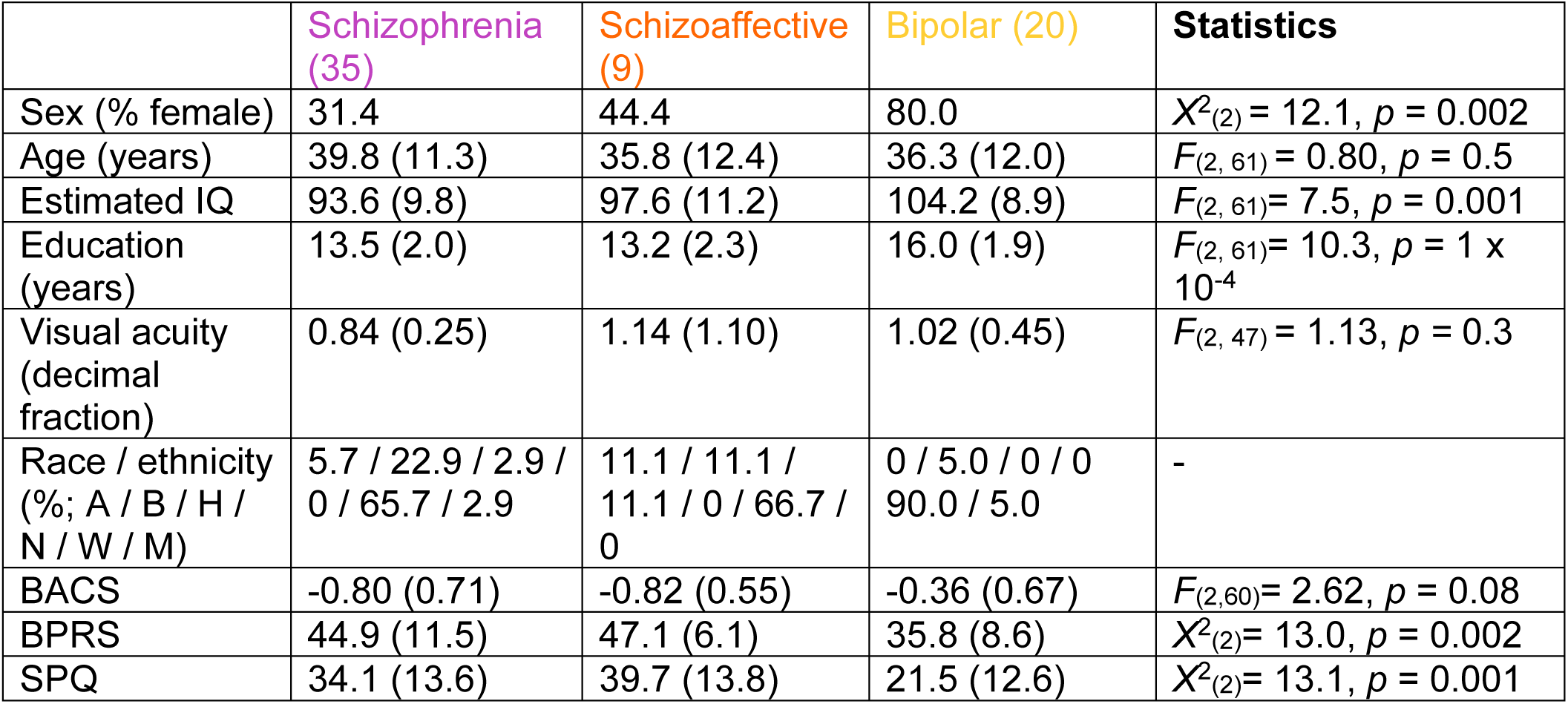
Participant demographics for diagnostic sub-groups of PwPP. Data are presented as mean (standard deviation), unless otherwise noted. Participant diagnoses were made using the Structured Clinical Interview for DSM-IV-TR disorders (SCID; First et al., 2002). Sex is presented in % female at birth. Age is presented in years. Estimated IQ was determined with the Wechsler Adult Intelligence Scale (WAIS-IV; Petermann & Wechsler, 2012). Racial and ethnic designations (as defined by the National Institute of Health) are abbreviated as follows: A = Asian or Pacific Islander, B = Black (not of Hispanic origin), H = Hispanic, N = Native American or Alaskan Native, W = White (not of Hispanic origin), M = More than 1 race or ethnicity, or other. BACS = Brief Assessment of Cognition in Schizophrenia, Z-score (Keefe, 2008), BPRS = Brief Psychiatric Rating Scale Total Score (Ventura et al., 2000), SPQ = Schizotypal Personality Questionnaire (Raine, 1991). Data collected at repeat scans were not included in this table. The test statistic and p value for differences across all 3 groups are shown in the statistics column. Non-parametric statistics (Kruskal-Wallis tests, X^2^ values) were used in place of ANOVAs in cases where data were not normally distributed and / or variance across groups was not homogeneous.

**Supplemental Table 3:**
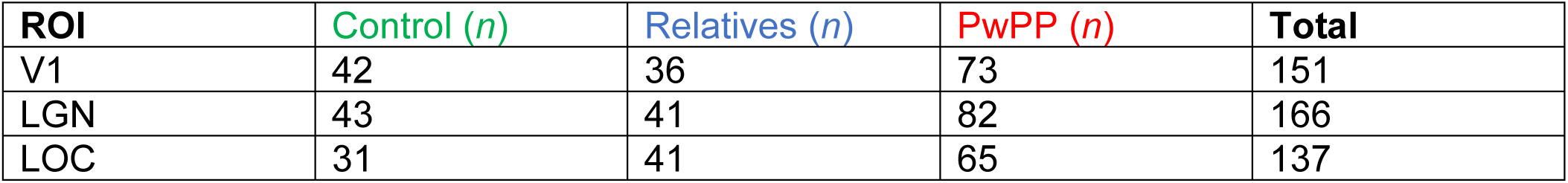
Number of ROIs available for each group. ROIs were combined across hemispheres when both were available. Numbers reflect total number of datasets, not unique individuals.

**Supplemental Figure 1:**
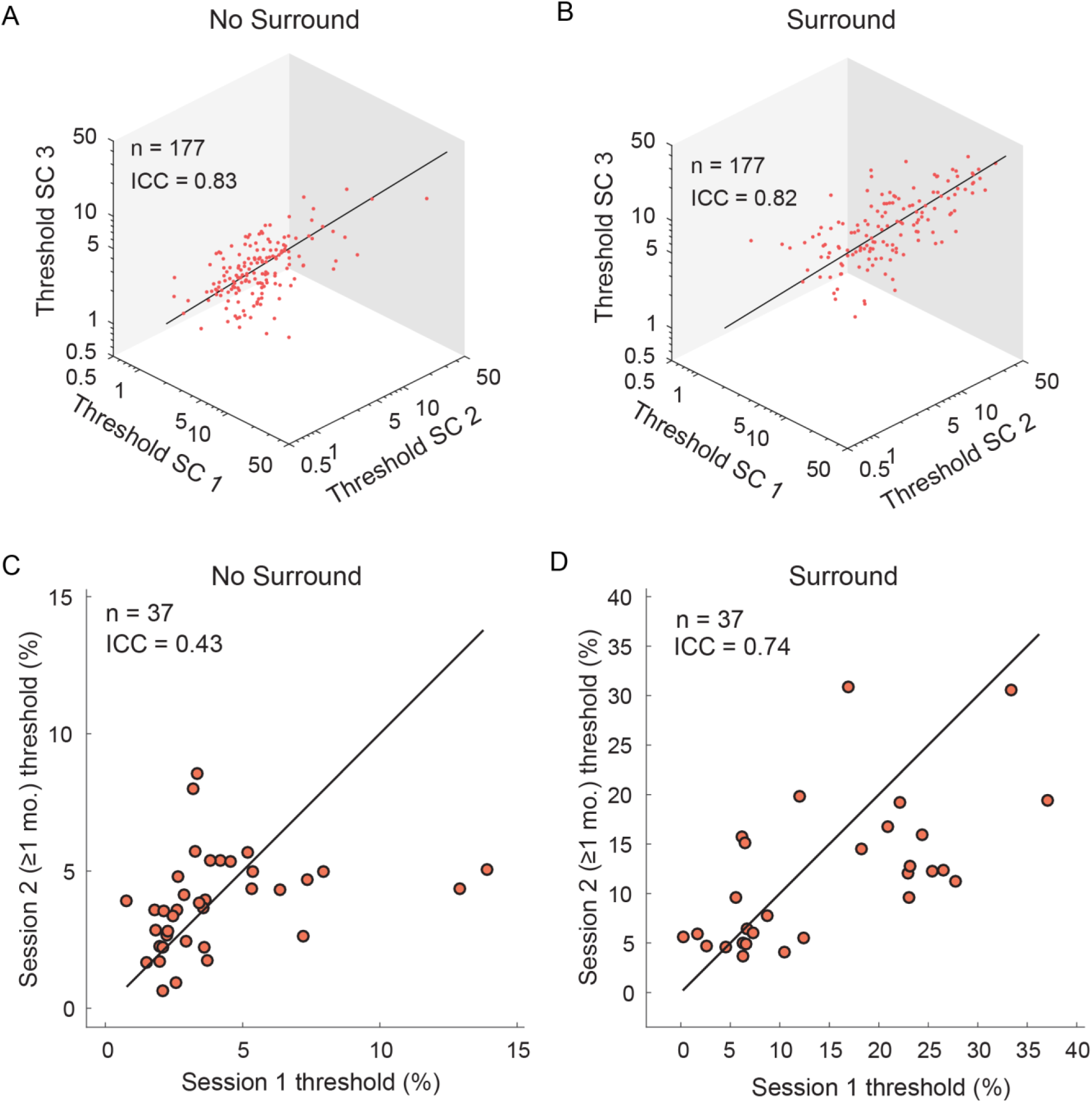
Test-retest reliability and longitudinal variability of contrast discrimination thresholds. A. Threshold estimates for No Surround are plotted for the three staircases (SC), showing high test-retest reliability within sessions (ICC = 0.83). B. Same, but for the Surround condition (ICC = 0.82). C. A subset of PwPP completed the experiments twice, with sessions separated by ≥1 month. This enabled us to quantify longitudinal variability for No Surround thresholds across sessions (ICC = 0.43). D. Same, but for Surround (ICC = 0.74). Black lines are unity.

#### Magnetic Resonance Spectroscopy (MRS)

MRS data were acquired from the medial occipital cortex using a stimulated echo acquisition mode (STEAM) sequence (TR = 5 s, TE = 8 ms, mixing time = 32 ms, hsinc pulse = 1.28 ms, volume-of-interest (VOI) size = 30 mm (left-right) x 18 mm (anterior-posterior) x 18 mm (superior-inferior), transmitter frequency = 3 ppm, 3D outer volume suppression interleaved with variable power and optimized relaxation delay (VAPOR) water suppression (Tkáč et al., 2001). 2048 complex data points were acquired with 6000 Hz spectral bandwidth. The chemical shift displacement error was 4% per ppm. We placed the VOI within the medial occipital lobe, superior to the cerebellar tentorium, posterior to the occipitoparietal junction, and anterior to the sagittal sinus. The position was adjusted as necessary to achieve a water linewidth of ≤ 15 Hz after shimming with FAST(EST) MAP.

The MRS data were processed in *matspec* toolbox in MATLAB, which included eddy current compensation, as well as frequency and phase correction. We quantified levels of metabolites within the VOI using LCModel (Provencher, 1993). We quantified concentrations for a total of 18 metabolites, though only GABA was used for the analyses in this manuscript. After correcting for differences in white matter, grey matter, and cerebrospinal fluid fractions within the VOI from each individual data set, metabolite levels were scaled relative to the unsuppressed water signal. MRS data that did not meet the following quality criteria were excluded: H_2_O line width ≤ 15 Hz, LCModel spectrum line width ≤ 5 Hz, and LCModel signal to noise ratio ≥ 40. Ten data sets (1 control, 4 relatives, 5 PwPP) were excluded, leaving 141 data sets (43 controls, 40 relatives, and 58 PwPP) for further analysis. For participants who completed repeat MRS scans, only the first was used for analysis. All spectroscopy and psychophysics data for a given participant were collected within 2 weeks. For all but one participant, they were acquired on the same day.

To determine whether greater concentrations of occipital GABA were associated with enhanced surround suppression, we applied Spearman’s ranked correlations between GABA measured in the occipital cortex and normalized suppression metrics. We first applied the correlation across all participant groups, then within each group. FDR corrections for multiple comparisons were applied to the three within-group correlations. We also tested for group differences in GABA levels with a non-parametric Kruskal-Wallis test.

### Supplemental Results

#### FMRI

Lateral geniculate nucleus (LGN) fMRI responses were higher in PwPP than controls, and responses increased slightly with contrast (see Main results). We did not find significant differences between LGN fMRI responses with and without surround (Supplemental Figure 2).

**Supplemental Figure 2:**
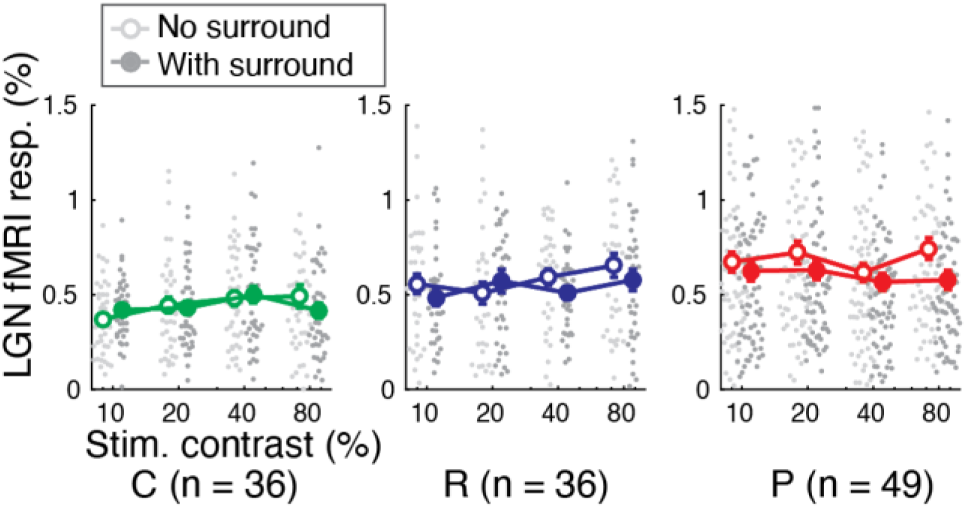
FMRI responses in LGN. Data from controls (C, green; n = 36), relatives (R, blue; n = 36), and PwPP (P, red; n = 49) for target gratings with (closed) and without 100% contrast surrounding gratings (open) at four pedestal contrast levels (10%, 20%, 40%, 80%; x axis). Light (No Surround) and dark grey (Surround) dots show individual data points, larger circles show group means. Error bars are standard error of the mean. Data from repeat scanning sessions are included for 5 controls and 25 PwPP (10 SZ, 11 BP, 4 SCA).

As fMRI suppression in LOC for Surround vs. No Surround was weaker in both PwPP and relatives vs. controls, we examined whether LOC suppression was associated with differences in measures related to psychiatric symptoms across participants. Specifically, we used Spearman’s correlations with BPRS total scores, BACS composite *Z* scores, SPQ total scores, and SGI total scores. Weaker LOC suppression (higher values) was associated with higher psychiatric symptom levels (higher BPRS total score; *r*_94_ = 0.22, *p_uncorr._* = 0.035; *p_FDR_* = 0.070; Supplemental Figure 3A) and poorer cognition (lower BACS *Z* score; *r*_98_ = −0.05, *p_uncorr._* = 0.035; *p_FDR_* = 0.104; Supplemental Figure 3B) across all participants, but these did not survive correction for multiple comparisons. As correlations within PwPP alone were not significant (uncorrected *p* values > 0.3), these relationships may have been driven by overall group differences in LOC suppression and symptom levels. Other correlations were not significant (Spearman’s correlations, SPQ: *r*_98_ = 0.06, *p_uncorr._* = 0.6; SGI: *r*_98_ = 0.19, *p_uncorr._* = 0.056, not shown).

**Supplemental Figure 3.**
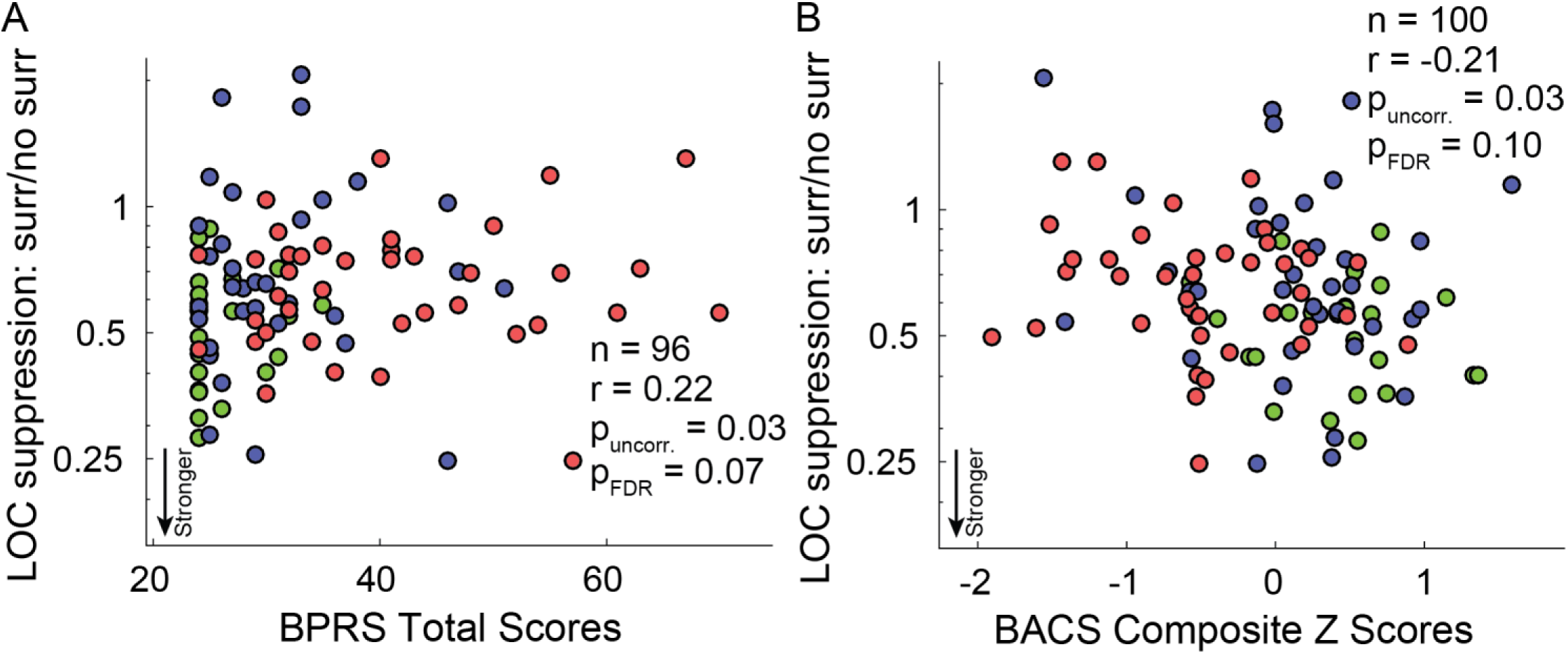
Correlation of LOC suppression with symptom measures. A) Suppression of the LOC fMRI response for Surround / No Surround conditions (y-axis) and BPRS total scores (x-axis) are plotted for all groups (Controls, green, n = 23; Relatives, blue, n = 34; PwPP, n = 39). B) Same, but for BACS composite Z scores (x-axis; Controls, green, n = 24; Relatives, blue, n = 36; PwPP, n = 40).

**Supplemental Figure 4:**
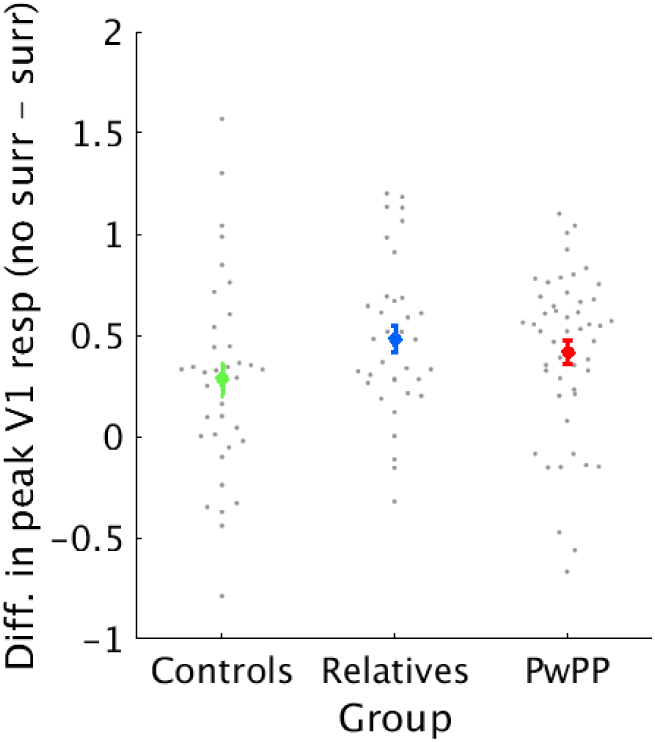
Difference in peak V1 responses, used in our decoding analysis. Difference between No Surround and Surround V1 fMRI responses (average of the peak response, from 7-9 s after stimulus onset). Gray dots show individual data points, colored dots show group means for controls (green), relatives (blue), and PwPP (red). Error bars are S.E.M. Differences in response for No Surround minus Surround tended to be positive, showing the expected surround suppression effect. Differences in responses were comparable across groups, suggesting that group differences in decoding accuracy (*Figure 6*) may not be explained by group differences in response magnitude.

#### Decoding

We examined decoding accuracy for V1 fMRI responses across all 8 conditions (4 contrasts, each with and without surrounding gratings) at each time point within the stimulus epoch starting 2 s before block onset and ending 6 s after block offset. As expected, decoding accuracy changed over time (ANOVA, *F*_17,139_ = 13.02, *p* = 9 × 10^−36^), with accuracy peaking near the end of the 9 s block. While there were no overall differences in decoding accuracy across groups, there was a significant interaction between group and time (*F*_34,2363_ = 1.54, *p* = 0.024). Given that the time-course of decoding accuracy varied across groups, we chose to focus on the time of the peak of the univariate V1 fMRI response (7-9 s after block onset). Across all participants, decoding accuracy during this peak response window was significantly greater than chance (i.e., higher than when the classifier was trained on randomized condition labels; two-sample *t*-test, *t*_141_ = −9.65, *p* = 3 × 10^−17^; Supplemental Figure 5).

**Supplemental Figure 5:**
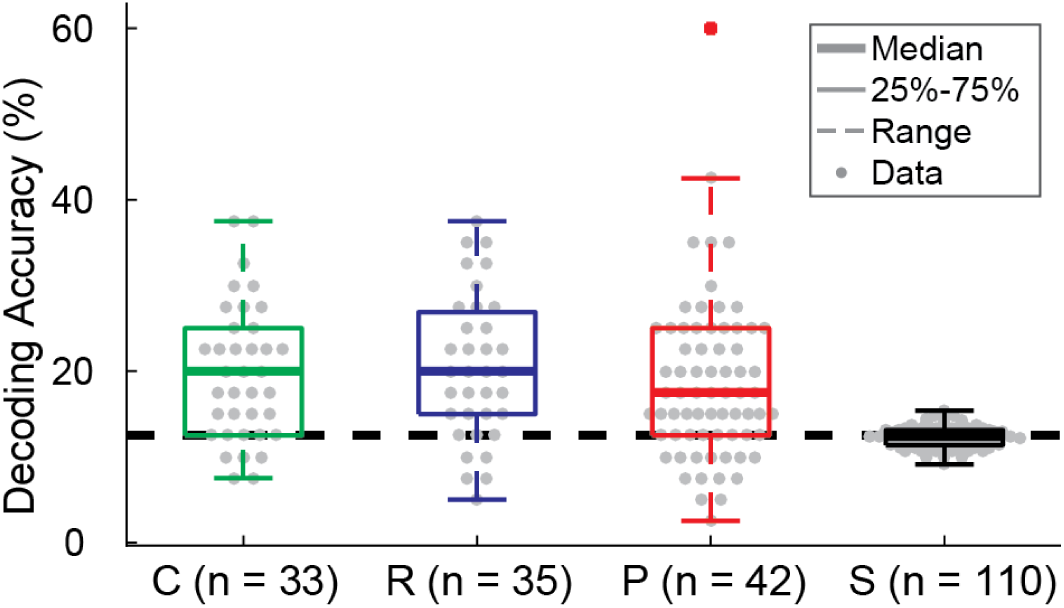
V1 fMRI decoding accuracy for all eight conditions. Decoding accuracy of fMRI values averaged across 7-9s—during the peak univariate response—for each group (Controls, C, n = 33, Relatives, R, n = 35, and PwPP, P, n = 42 unique individuals). Boxplots show median (thick line), 25%, and 75% quartiles. A dashed line shows 1.5x the interquartile range. Individual data are shown as grey circles for each group. For comparison, we show two depictions of chance. The dashed black line at 12.5% shows statistical chance, and a boxplot of decoding accuracy when the classifier was trained on data with randomly shuffled labels is shown in black (Shuffled, S, includes all participants, n = 110).

We also assessed whether multi-voxel patterns reflected differences in stimulus contrast (regardless of surround). To examine this, we trained an SVM classifier to decode stimulus contrast (i.e., collapsing across surround conditions). Across all participants, we found that decoding accuracy for contrast was significantly greater than chance (i.e., with shuffled contrast labels; paired *t*-test, *t*_141_ = −11.5, *p* = 7 × 10^−22^). However, we did not find significant group differences in contrast decoding accuracy in V1 (ANOVA, *F*_2,139 =_ 0.30, *p* = 0.7, Supplemental Figure 6).

**Supplemental Figure 6:**
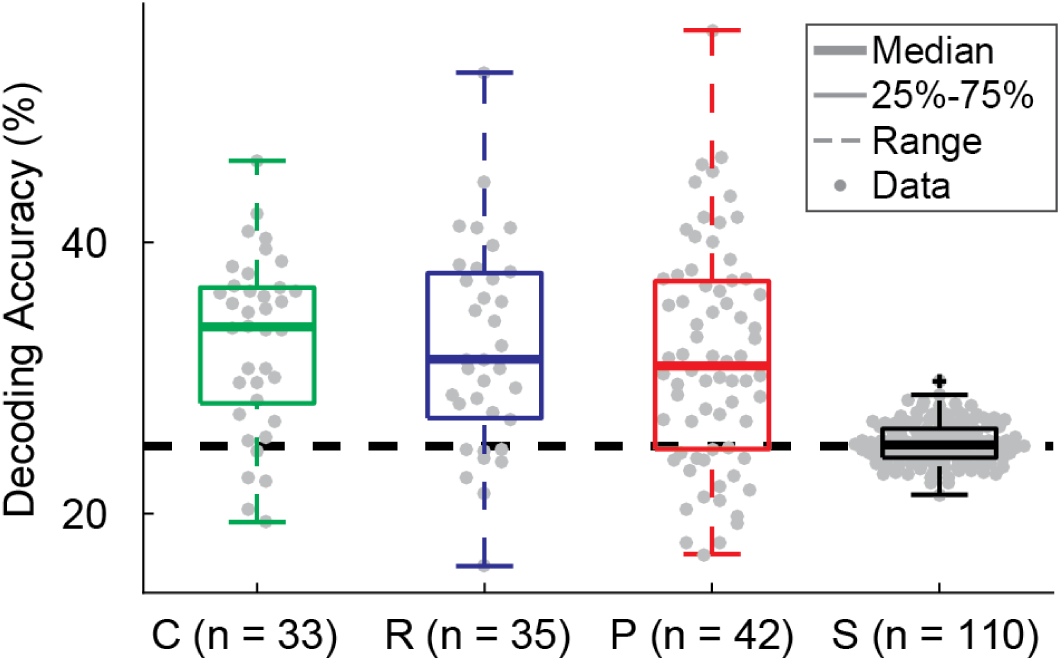
Decoding contrast in V1. Boxplots of decoding accuracy for each group (Controls, C, n = 33, Relatives, R, n = 35, PwPP, P, n = 42 unique individuals) separately. Individual scan decoding accuracies are plotted as grey dots. Two representations of chance are also plotted; a dashed line at 25% represents statistical chance, and a box plot of decoding accuracies when the classifier was trained with randomly shuffled labels is presented at the far right (Shuffled, S, all participants, n = 110).

Given the group differences in decoding accuracy for Surround vs. No Surround reported in the main text, we decided to further investigate whether there were any relationships between symptoms and surround decoding accuracy. We calculated Spearman’s correlations with BPRS total scores, BACS composite *Z* scores, SPQ total scores, and SGI total scores. Across all 3 participant groups, BPRS total scores were anti-correlated with surround decoding accuracy, but this did not survive correction for multiple comparisons (Spearman’s correlation, *r*_98_ = −0.21, *p_uncorr._* = 0.03, *p_FDR_* = 0.13; ^S^upplemental Figure ^7^). This relationship was not significant among PwPP alone (Spearman’s correlation, *r*_41_ = −0.12, *p_uncorr._* = 0.5), suggesting that the correlation across all participants may be explained by group differences in both decoding accuracy and BPRS scores. Other correlations were not significant (Spearman’s correlations, BACS: *r*_109_ = 0.1, *p_uncorr._* = 0.3; SPQ: *r*_110_ = −0.14, *p_uncorr._* = 0.15; SGI: *r*_110_ = −0.07, *p_uncorr._* = 0.5; not shown).

We also investigated whether we could reliably decode stimulus condition from the LGN fMRI response, as with V1 above. There were no significant effects of time or group on LGN fMRI decoding accuracy (ANOVA, *F* values < 1.2, *p* values > 0.2). As we saw no significant main effect of time in LGN, we did not conduct further analyses of the peak univariate response window. Finally, we also conducted decoding analyses for fMRI responses in LOC. As expected, there was a significant effect of time, where decoding accuracy increased following the onset of the stimulus (ANOVA, *F*_17,132_ = 9.62, *p* = 6 × 10^−25^). There was no main effect of group for decoding accuracy in LOC (ANOVA, *F*_2,132_ = 1.1, *p* = 0.3), nor was there a significant group by time interaction (*F*_34,2244_ = 0.79, *p* = 0.8). Given the lack of group differences, we did not examine decoding of the peak fMRI response window in LOC.

**Supplemental Figure 7:**
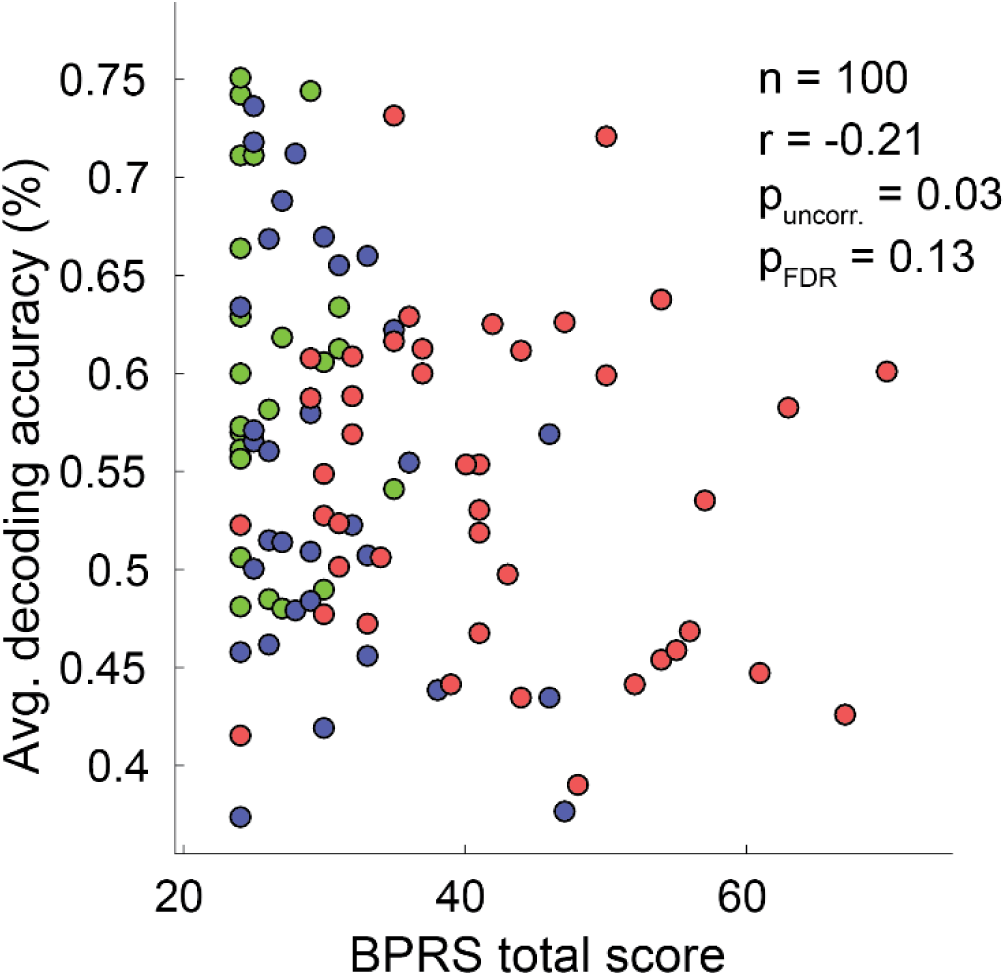
Correlation between V1 decoding accuracy for presence of surround vs. BPRS total scores. A) Decoding accuracy (y-axis) and individuals’ BPRS scores from the first visit (x-axis) are plotted for all groups (Controls, green, n = 25; Relatives, blue, n = 32; PwPP, n = 43).

#### MRS

We examined whether surround suppression was associated with the concentration of the inhibitory neurotransmitter GABA within visual cortex. Occipital GABA levels were acquired at rest using 7 T MRS. We did not find any significant group differences in GABA levels between controls, relatives, and PwPP (Kruskal Wallis, *Χ*^2^_2_ = 0.09 *p* = 1.0; Supplemental Figure 8A) nor any significant correlations between behavioral suppression indices and GABA levels within or across groups (all participants: Spearman’s correlation, *r*_117_ = −0.003, *p* = 1.0; each group separately: |*r* values| < 0.26, uncorrected *p* values > 0.07, FDR corrected *p* values > 0.2; Supplemental Figure 8B). Furthermore, we found no evidence for significant correlations between occipital GABA concentrations and V1 suppression indices within or across groups (Spearman’s correlation, *r*_108_ = −0.03, *p* = 0.8; Each group separately: (|*r* values| < 0.25, *p_uncorr_*. values > 0.15, *p_FDR_* values > 0.4; Supplemental Figure 8C). Thus, we did not find any evidence to support a direct link between GABA levels and suppression in our sample.

**Supplemental Figure 8:**
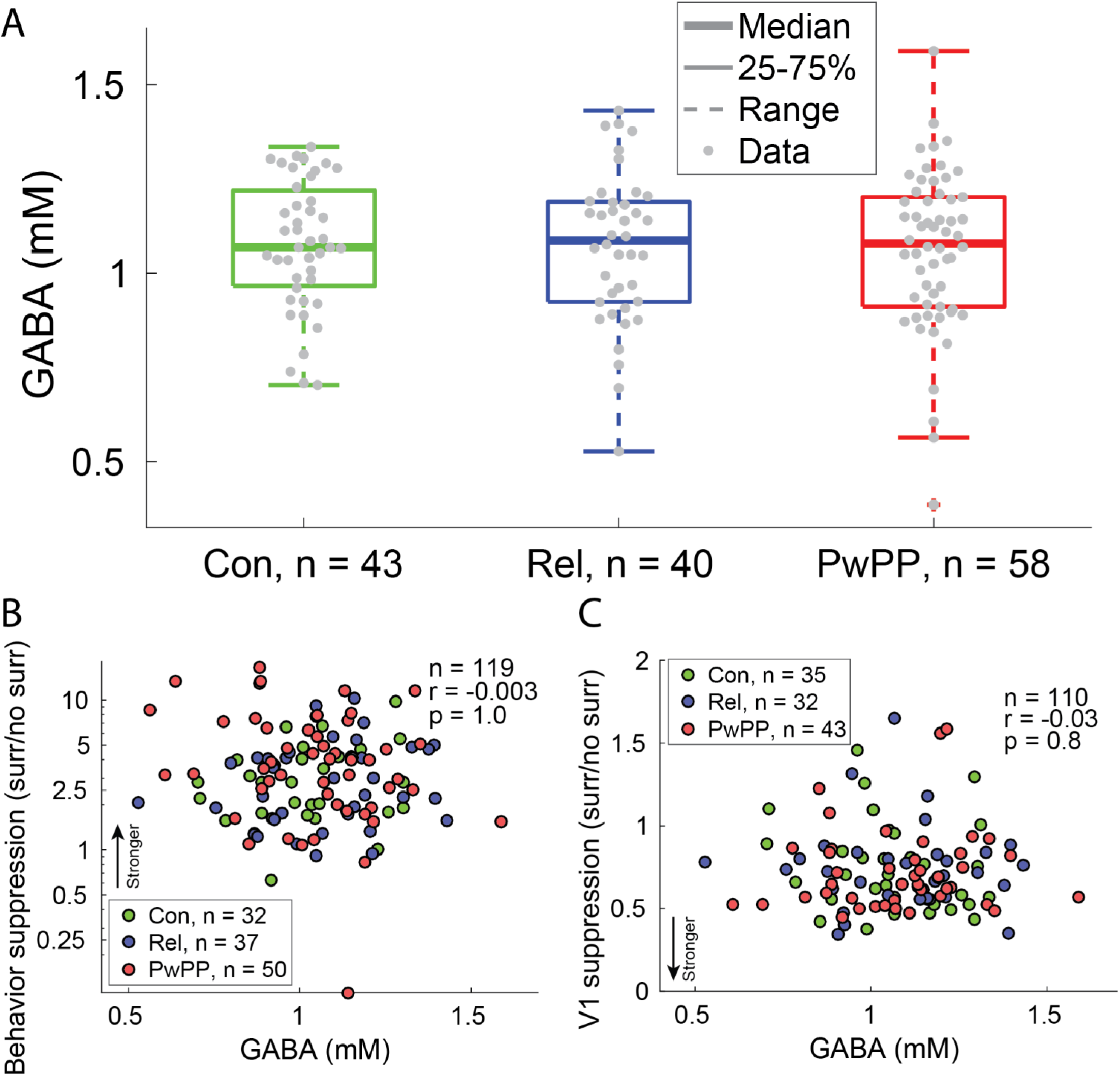
GABA levels measured with MR Spectroscopy do not differ between groups and do not correlate with surround suppression. A) GABA levels (mM) measured in occipital cortex are shown for each group (Controls, Con, n = 43; Relatives, Rel, n = 40; PwPP, n = 58). Individual data are plotted as grey dots. Box plots show the median (thick line), 25% to 75% interquartile range (box), 1.5x interquartile range (dashed line). B) Correlations between behavioral suppression metrics (y) and GABA concentration (x) are plotted for all participants. C) Correlations between V1 suppression metrics (y) and GABA concentration (x) are plotted for all participants. In both B) and C), the dot color indicates which group participants belonged to (green: Controls; blue: Relatives; red: PwPP). Correlations between occipital GABA levels and suppression metrics were not significant.

